# Fit notes associated with COVID-19 in 24 million patients’ primary care records: A cohort study in OpenSAFELY-TPP

**DOI:** 10.1101/2023.07.28.23293269

**Authors:** Andrea L Schaffer, Robin Y Park, John Tazare, Krishnan Bhaskaran, Brian MacKenna, Spiros Denaxas, Iain Dillingham, Sebastian CJ Bacon, Amir Mehrkar, Chris Bates, Ben Goldacre, Felix Greaves, John Macleod, The OpenSAFELY Collaborative, National Core Studies Collaborative, Laurie A Tomlinson, Alex J Walker

**Author notes:** Corresponding author: Dr Andrea Schaffer. ^Members of the collaboratives are listed in the Acknowledgements (The OpenSAFELY Collaborative, National Core Studies Collaborative).

## Abstract

**Background:** Fit notes (“sick notes”) are issued by general practitioners (GPs) when a person can’t work for health reasons and is an indication of the public health and economic burden for people recovering from COVID-19.

**Methods:** With NHS England approval, we used routine clinical data from >24 million patients to compare fit note incidence in people 18-64 years with and without evidence of COVID-19 in 2020, 2021 and 2022. We fit Cox regression models to estimate adjusted hazard ratios, overall and by time post-diagnosis and within demographic subgroups.

**Results:** We identified 365,421, 1,206,555 and 1,321,313 people with evidence of COVID-19 in 2020, 2021 and 2022. The fit note rate was 4.88 per 100 person-months (95%CI 4.83-4.93) in 2020, 2.66 (95%CI 2.64-2.67) in 2021, and 1.73 (95%CI 1.72-1.73) in 2022. Compared with the age, sex and region matched general population, the hazard ratio (HR) adjusted for demographics and clinical characteristics over the follow-up period was 4.07 (95%CI 4.02-4.12) in 2020 decreasing to 1.57 (95%CI 1.56-1.58) in 2022. The HR was highest in the first 30 days post-diagnosis in all years.

**Conclusions:** Despite likely underestimation of the fit note rate, we identified a considerable increase among people with COVID-19, even in an era when most people are vaccinated. Most fit notes are associated with the acute phase of the disease, but the increased risk several months post-diagnosis provides further evidence of the long-term impact.

**Evidence before this study:** We searched Pubmed from 1 March 2020 to 30 June 2023 using the following search terms: (“COVID-19” OR “SARS-CoV-2” OR “coronavirus”) AND (“United Kingdom” OR “England” OR “Britain” OR “Scotland” OR “Wales”) AND (“fit note” OR “sick note” OR “sick leave” OR “sickness absence”). We also searched the reference list of relevant articles. We included both peer-reviewed research studies and grey literature that quantified receipt of fit notes or sick leave during the COVID-19 pandemic.

We found two peer-reviewed studies and one briefing by an independent think tank. A study of 959,356 National Health Service (NHS) employees in England quantified receipt of non-COVID-19 related fit notes during the first wave of the pandemic. They found that the overall fit note rate was lower in 2020 compared with 2019. However, increases in the number of people receiving fit notes were observed for respiratory, infectious disease, and mental health conditions. The second study of 15,931 domiciliary care workers in Wales between Mar 2020 and Nov 2021 found that 15% had been issued a fit note over the study period. Fit notes were more common among women, people ≥45 years, and those with comorbidities. The briefing found that the percentage of sickness absence days taken by NHS employees was higher in 2022 (5.6%) compared with 2019 (4.3%), with a particular increase in absences due to mental health and infectious diseases. In 2022, 18% of sickness absence days were attributable to COVID-19.

**Added value of this study:** This study is the first to quantify changes in fit note rate since the start of the COVID-19 pandemic among people with a reported SARS-CoV-2 infection and how this compares with the general population in the UK. We found that people with evidence of SARS-CoV-2 infection had a higher fit note rate than the general population, even after adjusting for demographics and clinical characteristics. While this increased risk was greatest in 2020 (hazard ratio [HR] = 4.07, 95%CI 4.02-4.12), it continued to a lesser extent even into 2022 (HR = 1.57, 95%CI 1.56-1.58). The fit note rate was greatest in the first 30 days post-diagnosis, suggesting that most sick leave is associated with the acute phase. In subgroup analyses, the groups with the greatest relative increased risk changed over the years. People aged 18-24 years had a larger relative increased risk of fit notes (as measured by HR) in 2022 than 2021, when compared with the general population in each year. Additionally, while in 2020 and 2021 the HR increased along with lessening deprivation, this effect dissipated in 2022. In contrast, people hospitalised with COVID-19 were less likely to be issued a fit note than the pneumonia cohort, suggesting the long-term effects may be similar to comparable severe respiratory infections cases resulting in hospitalisation.

**Implications of all the available evidence:** While we have likely underestimated the fit note rate due to overcounting of people in the workforce and misclassification of COVID-19 status, we still identified a substantial increased risk of receiving a fit note in people with COVID-19 compared with the general population over all years, even after adjusting for demographics and a wide range of clinical characteristics. The increased risk persisted into 2022, in an era where most people are vaccinated and the severity of COVID-19 illness is lessened. Given the high infection rates still occurring, these findings provide evidence for a substantial impact of COVID-19 on productivity and further evidence of the long-term impacts of COVID-19.

## Background

In primary care, a doctor may issue a fit note (also called “sick note”) after the first seven days of sickness absence if the doctor assesses that the patient’s health affects their fitness for work. In 2021-22, over 11 million fit notes were issued in England(1). Long-term sickness absence from employment has negative consequences for the economy as well as individuals and can lead to widened health inequalities, financial insecurity, and reduced social participation.(2) Improving the health and productivity of the population and reducing welfare benefit claims are important policy objectives.(3,4)

The Office of National Statistics (ONS) estimated that 82% of the English population had been infected with SARS-CoV-2 by November 2022.(5,6) Additionally, an estimated 1.9 million people self-reported as experiencing long COVID symptoms in the UK in the four weeks prior to March 2023, with two-thirds experiencing symptoms for at least one year.(7) The prevalence of long-term symptoms was greatest in people aged 35-69 years, women, and people living in more deprived areas.(7) Previous UK research has also shown that receipt of fit notes varies by sociodemographics, with women and people in manual and service occupations more likely to be issued a fit note.(4) The most common reasons for receiving a fit note prior to the COVID-19 pandemic included mental health and musculoskeletal disorders.(1,8) To date, there has been limited research on fit notes issued to patients recovering from COVID-19 in England.

Given the risk of long-term symptoms following COVID-19 (“long COVID”), it is of public health interest to quantify the impact of SARS-CoV-2 infection and recovery on the workforce. However, long COVID is heterogeneous and difficult to define.(9,10) Furthermore, coding of long COVID in primary care is very low,(11) and cannot be relied on to identify people suffering from persistent symptoms. Therefore, our objectives were to: 1) describe the demographic and clinical characteristics of people given a fit note following a documented SARS-CoV-2 infection or COVID-19 diagnosis; 2) determine how the fit note rate varies over time post-diagnosis; and 3) quantify the difference in fit note rate in people with SARS-CoV-2 infection or COVID-19 diagnosis compared with the general population and people hospitalised with pneumonia.

## Methods

### Study design

We conducted an observational cohort study using general practice primary care electronic health record (EHR) data from primary care practices in England.

### Data source and data sharing

We used primary care data from approximately 40% of the English population currently registered with GP surgeries using TPP SystmOne software. All data were linked, stored and analysed securely using the OpenSAFELY platform, https://www.opensafely.org/, as part of the NHS England OpenSAFELY COVID-19 service. Data include pseudonymised data such as coded diagnoses, medications and physiological parameters. No free text data are included. All code is shared openly for review and re-use under MIT open license (https://github.com/opensafely/long-covid-sick-notes). Similarly, pseudonymised datasets including ONS registered deaths, hospital episode statistics (HES), and Second Generation Surveillance System (SGSS) COVID-19 test results are securely provided to TPP and linked to primary care data. Detailed pseudonymised patient data are potentially re-identifiable and not shared.

### Study population

We included adults 18-64 years registered with one GP for at least one year prior to their index date with information on age, sex, index of multiple deprivation (IMD) and the Sustainability and Transformation Partnership region (STP, an NHS administrative region). This age range was selected to represent people most likely to be in the workforce. From this source population, we identified three cohorts with recorded SARS-CoV-2 infection between 1 February and 30 November in each of 2020, 2021 and 2022 (hereafter referred to as the “COVID-19 cohorts”). They were identified through three routes: having a recorded positive test for SARS-CoV-2 based on SGSS data, which captures both polymerase chain reaction (PCR) and lateral flow tests (LFT); having a probable diagnostic code for COVID-19 in primary care records; or being hospitalised with a primary or secondary diagnosis for COVID-19 (ICD-10 codes U07.1 or U07.2) identified from HES. The index date was the earliest of these events. As comparators, we identified contemporary (2020, 2021, 2022) and one historical (2019) general population cohort who were frequency-matched on age, sex and STP to the COVID-19 cohorts. The 2019 general population cohort was matched with the 2020 COVID-19 cohort. For the comparator cohorts, the index date was randomly assigned and randomly distributed over the study period.

We also performed a secondary analysis among the subset of people hospitalised with COVID-19. Here, the index date was the date of admission. We compared these individuals with people hospitalised with pneumonia between 1 February 2019 and 30 November 2019. The pneumonia cohort was identified using the following ICD-10 codes identified in any diagnosis position: B01.2, B05.2, B20.6, B25.0, J10-J18, J85.1, U04. No contemporary pneumonia comparator cohort was included due to potential for misclassification with COVID-19. People could be included in multiple cohorts if they met the inclusion criteria.

### Study measures

#### Primary outcome

We identified the first recorded fit note over patient follow-up. Fit notes were identified using Clinical Terms Version 3 (CTV3) codes. People who were required to isolate beyond 7 days due to having or living with someone with symptoms of COVID-19 would be issued an isolation note, rather than a fit note. Isolation notes did not require contact with a GP, and are not counted. Follow-up was censored at the end of the study period (30 November), death, or deregistration from their GP practice. The comparator cohorts were also censored if they had a recorded positive SARS-CoV-2 test or COVID-19 diagnosis.

#### Demographics and clinical characteristics

We characterised the demographic and clinical characteristics of patients who received a fit note. Measured demographic variables included age (18-24, 25-34, 35-44, 45-54, 55-64 years), sex (male, female), GP practice region (East Midlands, East, London, North East, North West, South East, South West, West Midlands, Yorkshire and The Humber), ethnicity (White, Asian and British Asian, Black, Other, Unknown), and Indices of Multiple Deprivation (IMD) quintiles.

Clinical variables included indicators for pre-existing conditions that may impact the severity of COVID-19, specifically: asthma, cancer (haematological, lung, other), chronic cardiac disease, chronic liver disease, chronic respiratory disease (excluding asthma), diabetes, asplenia, human immunodeficiency virus (HIV) infection, hypertension, obesity, organ transplant, neurological conditions, other permanent immunodeficiency (excluding HIV), smoking status (current, former or never), and autoimmune conditions (rheumatoid arthritis, systemic lupus erythematosus (SLE), psoriasis). Obesity and smoking status were identified using the most recent recorded information, while the remainder were identified at any time prior to the index date.

### Statistical methods

#### Rate of first fit note

For each of the eleven cohorts (three COVID-19 cohorts [2020, 2021, 2022], four matched general population cohorts [2019, 2020, 2021, 2022], three hospitalised COVID-19 cohorts [2020, 2021, 2022] and one hospitalised pneumonia cohort [2019]), we reported the first fit note rate per 100 person months overall and stratified by demographics (age, sex, ethnicity, region, IMD quintile). To prevent disclosure, all counts in this manuscript are rounded to the nearest 7.

#### Cox regression

We used Cox regression models to estimate hazard ratios (HRs) and 95%Confidence intervals (CIs) comparing the first fit note rate between the COVID-19 cohorts and the comparator cohorts. All of the COVID-19 cohorts were compared with their contemporary general population cohort as well as the historical 2019 general population cohort. The hospitalised COVID-19 cohorts were compared with the hospitalised pneumonia cohort (2019) only.

We investigated crude univariable and two covariate-adjusted models: (1) adjusted for age (cubic splines with 4 knots) and sex; (2) adjusted for demographics and all clinical characteristics described above. While the same people can contribute person-time to multiple exposure groups, these periods are non-overlapping, so we applied robust standard errors.

To determine if the relative differences in fit note occurrence changed over time, we re-estimated crude and adjusted HRs and 95%CIs censoring follow-up at 30, 90, and 150 days, and examined whether HRs changed according to the duration of follow-up. We chose to use this approach instead of estimating period-specific HRs in order to avoid selection bias due to depletion of susceptibles.(12) Similarly, to explore the impact within demographic categories, we estimated crude and adjusted HRs and 95%CIs stratified by age group, sex, ethnicity, IMD quintile, and region.

### Software and reproducibility

Data management was performed using Python 3.8, with analyses carried out using Stata 16.1, R 4.3.0, and Python. Code for data management and analysis as well as all codelists used in this study are available online: https://github.com/opensafely/long-covid-sick-notes.

### Patient and public involvement

We have a publicly available website https://opensafely.org/ through which we invite any patient or member of the public to contact us regarding this study or the broader OpenSAFELY project.

## Results

### Study population description

We identified 365,421 people with a recorded positive SARS-CoV-2 test or COVID-19 diagnosis in 2020, of which 22,015 (6.0%) were hospitalised; 1,206,555 people in 2021 (30,205 [2.5%] hospitalised); and 1,321,313 people in 2022 (34,692 [2.6%] hospitalised) (**Table 1**). The matched general population cohorts included 3.1 million people in 2019, 3.4 million in 2020, 4.6 million in 2021, and 4.8 million in 2022. For comparison with COVID-19 patients who were hospitalised, we identified 29,673 patients hospitalised with pneumonia in 2019. The majority of people in the COVID-19 cohorts were identified via SGSS testing (88.8% in 2020, 97.8% in 2021 and 96.5% in 2022) (**Supplementary Figure 1**).

**Table 1.** Demographic and clinical characteristics of all cohorts, 1 Feb - 30 Nov of each year. All counts rounded to nearest 7.

|  | 2019 | 2020 |  | 2021 |  | 2022 |  |
| --- | --- | --- | --- | --- | --- | --- | --- |
|  | General population* | COVID-19 cohort | General population^ | COVID-19 cohort | General population^ | COVID-19 cohort | General population^ |
|  | n (%) | n (%) | n (%) | n (%) | n (%) | n (%) | n (%) |
| **Total** | 3,140,326 (100.0) | 365,421 (100.0) | 3,439,534 (100.0) | 1,206,555 (100.0) | 4,571,469 (100.0) | 1,321,313 (100.0) | 4,818,870 (100.0) |
| **Age** |  |  |  |  |  |  |  |
| 18-24 y | 496,825 (15.8) | 61,355 (16.8) | 545,286 (15.9) | 196,616 (16.3) | 699,993 (15.3) | 111,342 (8.4) | 375,704 (7.8) |
| 25-34 y | 663,495 (21.1) | 77,686 (21.3) | 726,712 (21.1) | 264,159 (21.9) | 989,506 (21.6) | 258,314 (19.5) | 915,152 (19.0) |
| 35-44 y | 644,945 (20.5) | 74,767 (20.5) | 706,692 (20.5) | 294,238 (24.4) | 1,116,283 (24.4) | 312,732 (23.7) | 1,142,764 (23.7) |
| 45-54 y | 719,663 (22.9) | 83,832 (22.9) | 787,465 (22.9) | 271,418 (22.5) | 1,042,601 (22.8) | 328,510 (24.9) | 1,197,693 (24.9) |
| 55-64 y | 615,405 (19.6) | 67,781 (18.5) | 673,372 (19.6) | 180,117 (14.9) | 723,093 (15.8) | 310,422 (23.5) | 1,187,557 (24.6) |
| **Sex** |  |  |  |  |  |  |  |
| Female | 1,732,654 (55.2) | 201,824 (55.2) | 1,898,351 (55.2) | 645,666 (53.5) | 2,447,200 (53.5) | 823,438 (62.3) | 3,001,922 (62.3) |
| Male | 1,407,672 (44.8) | 163,597 (44.8) | 1,541,183 (44.8) | 560,889 (46.5) | 2,124,269 (46.5) | 497,875 (37.7) | 1,816,948 (37.7) |
| **Ethnicity** |  |  |  |  |  |  |  |
| White | 1,914,458 (61.0) | 213,185 (58.3) | 2,109,478 (61.3) | 788,249 (65.3) | 2,866,892 (62.7) | 967,225 (73.2) | 3,287,305 (68.2) |
| Asian or Asian British | 225,316 (7.2) | 45,101 (12.3) | 254,884 (7.4) | 68,915 (5.7) | 315,854 (6.9) | 52,101 (3.9) | 300,167 (6.2) |
| Black | 67,319 (2.1) | 8099 (2.2) | 76,622 (2.2) | 21,399 (1.8) | 103,117 (2.3) | 18,340 (1.4) | 105,322 (2.2) |
| Mixed | 34,748 (1.1) | 4298 (1.2) | 39,872 (1.2) | 13,937 (1.2) | 56,644 (1.2) | 14,077 (1.1) | 58,527 (1.2) |
| Other | 60,935 (1.9) | 5502 (1.5) | 70,427 (2.0) | 14,910 (1.2) | 102,585 (2.2) | 19,425 (1.5) | 105,007 (2.2) |
| Unknown | 837,550 (26.7) | 89,236 (24.4) | 888,265 (25.8) | 299,145 (24.8) | 1,126,384 (24.6) | 250,152 (18.9) | 962,542 (20.0) |
| **IMD** |  |  |  |  |  |  |  |
| 1 (most deprived) | 795,739 (25.3) | 98,399 (26.9) | 877,583 (25.5) | 257,992 (21.4) | 1,027,467 (22.5) | 212,044 (16.0) | 929,649 (19.3) |
| 2 | 630,189 (20.1) | 76,874 (21.0) | 691,131 (20.1) | 243,334 (20.2) | 926,135 (20.3) | 251,692 (19.0) | 968,303 (20.1) |
| 3 | 605,759 (19.3) | 68,887 (18.9) | 661,150 (19.2) | 248,696 (20.6) | 935,067 (20.5) | 292,131 (22.1) | 1,047,914 (21.7) |
| 4 | 57,9614 (18.5) | 64,064 (17.5) | 633,059 (18.4) | 235,473 (19.5) | 877,170 (19.2) | 286,538 (21.7) | 975,576 (20.2) |
| 5 (least deprived) | 52,9025 (16.8) | 57,190 (15.7) | 576,618 (16.8) | 221,067 (18.3) | 805,623 (17.6) | 278,915 (21.1) | 897,421 (18.6) |
| **Region** |  |  |  |  |  |  |  |
| East | 432,551 (13.8) | 50,246 (13.8) | 473,928 (13.8) | 240,170 (19.9) | 910,567 (19.9) | 327,096 (24.8) | 1,184,981 (24.6) |
| East Midlands | 645,218 (20.5) | 75,040 (20.5) | 706,370 (20.5) | 227,577 (18.9) | 863,884 (18.9) | 222,397 (16.8) | 814,919 (16.9) |
| London | 145,264 (4.6) | 17,073 (4.7) | 158,844 (4.6) | 56,693 (4.7) | 210,623 (4.6) | 71,113 (5.4) | 255,997 (5.3) |
| North East | 257,985 (8.2) | 29,988 (8.2) | 282,198 (8.2) | 71,169 (5.9) | 270,284 (5.9) | 59,059 (4.5) | 216,216 (4.5) |
| North West | 426,615 (13.6) | 49,623 (13.6) | 467,383 (13.6) | 129,934 (10.8) | 494,144 (10.8) | 108,857 (8.2) | 399,168 (8.3) |
| South East | 101,976 (3.2) | 11,942 (3.3) | 111,986 (3.3) | 70,763 (5.9) | 266,098 (5.8) | 100,114 (7.6) | 363,867 (7.6) |
| South West | 220,759 (7.0) | 25,641 (7.0) | 241,500 (7.0) | 160,678 (13.3) | 608,790 (13.3) | 227,731 (17.2) | 833,742 (17.3) |
| West Midlands | 175,581 (5.6) | 20,559 (5.6) | 192,836 (5.6) | 51,737 (4.3) | 194,593 (4.3) | 40,880 (3.1) | 147,791 (3.1) |
| Yorkshire & The Humber | 734,370 (23.4) | 85,309 (23.3) | 804,496 (23.4) | 197,827 (16.4) | 752,486 (16.5) | 164,073 (12.4) | 602,175 (12.5) |
| **Health conditions** |  |  |  |  |  |  |  |
| Obesity | 681,422 (21.7) | 96,712 (26.5) | 748,398 (21.8) | 276,759 (22.9) | 957,194 (20.9) | 350,777 (26.5) | 1,137,710 (23.6) |
| Hypertension | 325,969 (10.4) | 43,610 (11.9) | 353,416 (10.3) | 112,147 (9.3) | 411,803 (9.0) | 164,367 (12.4) | 564,823 (11.7) |
| Diabetes | 184,450 (5.9) | 30,590 (8.4) | 210,532 (6.1) | 70,861 (5.9) | 262,906 (5.8) | 100,541 (7.6) | 358,449 (7.4) |
| Asthma | 538,657 (17.2) | 68,208 (18.7) | 595,763 (17.3) | 236,439 (19.6) | 807,618 (17.7) | 279,944 (21.2) | 852,586 (17.7) |
| Other chronic respiratory disease | 55,265 (1.8) | 6650 (1.8) | 59,773 (1.7) | 15,932 (1.3) | 67,361 (1.5) | 27,083 (2.0) | 90,755 (1.9) |
| Chronic cardiac disease | 79,667 (2.5) | 11,039 (3.0) | 86,933 (2.5) | 26,320 (2.2) | 100,527 (2.2) | 38,906 (2.9) | 127,764 (2.7) |
| Lung cancer | 1442 (0.0) | 280 (0.1) | 1610 (0.0) | 483 (0.0) | 1764 (0.0) | 1050 (0.1) | 2730 (0.1) |
| Haematological cancer | 9212 (0.3) | 1316 (0.4) | 10,101 (0.3) | 3444 (0.3) | 12,964 (0.3) | 7854 (0.6) | 16,142 (0.3) |
| Other cancer | 63,987 (2.0) | 7798 (2.1) | 69,923 (2.0) | 22,351 (1.9) | 84,742 (1.9) | 39,599 (3.0) | 125,146 (2.6) |
| Chronic liver disease | 14,672 (0.5) | 2093 (0.6) | 16,380 (0.5) | 4452 (0.4) | 20,860 (0.5) | 7637 (0.6) | 26,110 (0.5) |
| Other neurological disease | 21,945 (0.7) | 2499 (0.7) | 23,730 (0.7) | 6678 (0.6) | 29,827 (0.7) | 14,364 (1.1) | 35,924 (0.7) |
| Organ transplant | 3339 (0.1) | 679 (0.2) | 3591 (0.1) | 1554 (0.1) | 4494 (0.1) | 3885 (0.3) | 5257 (0.1) |
| Asplenia | 3416 (0.1) | 427 (0.1) | 3675 (0.1) | 1127 (0.1) | 4662 (0.1) | 2191 (0.2) | 5446 (0.1) |
| HIV | 5404 (0.2) | 714 (0.2) | 6027 (0.2) | 1799 (0.1) | 7973 (0.2) | 3304 (0.3) | 8799 (0.2) |
| Other permanent immunodeficiency | 2338 (0.1) | 336 (0.1) | 2730 (0.1) | 1071 (0.1) | 3689 (0.1) | 1869 (0.1) | 4158 (0.1) |
| Rheumatoid arthritis / SLE / psoriasis | 137,536 (4.4) | 16,982 (4.6) | 150,787 (4.4) | 56,203 (4.7) | 193,893 (4.2) | 71,022 (5.4) | 229,537 (4.8) |
| **Smoking status** |  |  |  |  |  |  |  |
| Current | 637,581 (20.3) | 47,915 (13.1) | 684,236 (19.9) | 194,327 (16.1) | 895,923 (19.6) | 192,381 (14.6) | 910,245 (18.9) |
| Former | 802,263 (25.5) | 103,810 (28.4) | 880,390 (25.6) | 358,526 (29.7) | 1,189,923 (26.0) | 426,041 (32.2) | 1,426,432 (29.6) |
| Never | 1,536,297 (48.9) | 194,747 (53.3) | 1,672,286 (48.6) | 581,210 (48.2) | 2,185,834 (47.8) | 661,794 (50.1) | 2,280,509 (47.3) |
| Missing | 164,185 (5.2) | 18,942 (5.2) | 202,622 (5.9) | 72,485 (6.0) | 299,789 (6.6) | 41,097 (3.1) | 201,684 (4.2) |

\*age, sex and STP frequency matched with 2020 COVID-19 cohort; ^age, sex and STP frequency matched with contemporary COVID-19 cohort; HIV = human immunodeficiency virus; SLE = systemic lupus erythematosus

Compared with 2020 and 2021, the 2022 COVID-19 cohort was older and more likely to be female, White ethnicity, and in less deprived IMD quintiles. Hospitalised COVID-19 cohorts tended to be older, and were more likely to be male, non-White ethnicity, and in the most deprived IMD quintile compared with the overall COVID-19 cohorts (**Supplementary Table 1**). When compared with the pneumonia cohort, hospitalised COVID-19 patients were more likely to be obese,less likely to be current smokers, and had lower rates of many chronic health conditions.

### Overall fit note rates

A total of 34,377 (9.4%), 102,949 (8.5%) and 152,859 (11.6%) of people in the COVID-19 cohorts were issued fit notes during follow-up in 2020, 2021 and 2022 (**Supplementary Table 2**). However, after taking into account differences in follow-up time (**Supplementary Figure 2**), the fit note rate among the COVID-19 cohorts decreased over time, from 4.88 per 100 person-months in 2020 (95%CI 4.83-4.93), to 2.66 (95%CI 2.64-2.67) in 2021 and 1.73 (95%CI 1.72-1.73) in 2022 (**Table 2**).

**Table 2.** Rate of first fit note per 100 person-months for each cohort by demographics.

|  | 2019 | 2020 |  | 2021 |  | 2022 |  |
| --- | --- | --- | --- | --- | --- | --- | --- |
|  | General population^ | COVID-19 cohort | General population^ | COVID-19 cohort | General population^ | COVID-19 cohort | General population^ |
|  | Rate per 100 person-months (95%CI) | Rate per 100 person-months (95%CI) | Rate per 100 person-months (95%CI) | Rate per 100 person-months (95%CI) | Rate per 100 person-months (95%CI) | Rate per 100 person-months (95%CI) | Rate per 100 person-months (95%CI) |
| **Total** | 1.21 (1.21 - 1.21) | 4.88 (4.83 - 4.93) | 0.93 (0.93 - 0.93) | 2.66 (2.64 - 2.67) | 1.23 (1.23 - 1.23) | 1.73 (1.72 - 1.73) | 1.23 (1.23 - 1.23) |
| **Age** |  |  |  |  |  |  |  |
| 18-24 y | 0.98 (0.97 - 0.99) | 1.58 (1.50 - 1.65) | 0.66 (0.65 - 0.66) | 1.25 (1.23 - 1.28) | 0.96 (0.95 - 0.98) | 1.30 (1.28 - 1.33) | 0.91 (0.90 - 0.92) |
| 25-34 y | 1.18 (1.16 - 1.19) | 3.51 (3.42 - 3.61) | 0.86 (0.85 - 0.87) | 2.16 (2.13 - 2.19) | 1.17 (1.16 - 1.18) | 1.62 (1.60 - 1.64) | 1.14 (1.13 - 1.15) |
| 35-44 y | 1.19 (1.18 - 1.20) | 5.19 (5.07 - 5.31) | 0.91 (0.90 - 0.92) | 2.95 (2.92 - 2.99) | 1.24 (1.23 - 1.25) | 1.65 (1.63 - 1.66) | 1.19 (1.18 - 1.20) |
| 45-54 y | 1.35 (1.34 - 1.37) | 6.53 (6.40 - 6.65) | 1.08 (1.07 - 1.09) | 3.60 (3.55 - 3.64) | 1.38 (1.37 - 1.39) | 1.91 (1.90 - 1.93) | 1.35 (1.34 - 1.36) |
| 55-64 y | 1.30 (1.28 - 1.31) | 6.62 (6.48 - 6.75) | 1.09 (1.07 - 1.10) | 3.79 (3.74 - 3.85) | 1.33 (1.32 - 1.35) | 1.87 (1.85 - 1.89) | 1.32 (1.32 - 1.33) |
| **Sex** |  |  |  |  |  |  |  |
| Female | 1.39 (1.38 - 1.40) | 5.88 (5.80 - 5.95) | 1.11 (1.10 - 1.11) | 3.25 (3.22 - 3.27) | 1.46 (1.46 - 1.47) | 1.96 (1.95 - 1.97) | 1.41 (1.41 - 1.42) |
| Male | 0.99 (0.98 - 1.00) | 3.61 (3.54 - 3.68) | 0.72 (0.71 - 0.72) | 2.03 (2.01 - 2.05) | 0.96 (0.95 - 0.97) | 1.35 (1.34 - 1.37) | 0.94 (0.93 - 0.94) |
| **Ethnicity** |  |  |  |  |  |  |  |
| White | 1.30 (1.30 - 1.31) | 5.17 (5.10 - 5.24) | 1.00 (0.99 - 1.00) | 2.79 (2.77 - 2.81) | 1.31 (1.31 - 1.32) | 1.73 (1.72 - 1.74) | 1.29 (1.28 - 1.29) |
| Mixed | 1.03 (1.01 - 1.04) | 4.83 (4.69 - 4.97) | 0.86 (0.84 - 0.87) | 3.11 (3.05 - 3.18) | 1.10 (1.08 - 1.12) | 2.07 (2.02 - 2.12) | 1.16 (1.14 - 1.18) |
| Asian / Asian British | 1.24 (1.20 - 1.28) | 5.51 (5.18 - 5.84) | 0.91 (0.88 - 0.94) | 3.47 (3.34 - 3.61) | 1.29 (1.25 - 1.32) | 2.22 (2.14 - 2.31) | 1.42 (1.38 - 1.45) |
| Black | 1.22 (1.16 - 1.27) | 4.55 (4.11 - 4.98) | 0.92 (0.88 - 0.96) | 2.65 (2.51 - 2.80) | 1.19 (1.15 - 1.23) | 1.74 (1.66 - 1.83) | 1.25 (1.21 - 1.29) |
| Other | 0.62 (0.59 - 0.65) | 3.91 (3.56 - 4.25) | 0.47 (0.44 - 0.49) | 2.31 (2.18 - 2.44) | 0.66 (0.64 - 0.69) | 1.30 (1.24 - 1.37) | 0.75 (0.72 - 0.77) |
| Unknown | 1.09 (1.08 - 1.10) | 4.23 (4.13 - 4.33) | 0.83 (0.82 - 0.84) | 2.19 (2.16 - 2.22) | 1.10 (1.09 - 1.11) | 1.62 (1.60 - 1.64) | 1.09 (1.08 - 1.10) |
| **Region** |  |  |  |  |  |  |  |
| East Midlands | 1.05 (1.03 - 1.06) | 4.06 (3.94 - 4.17) | 0.76 (0.75 - 0.77) | 2.39 (2.36 - 2.43) | 1.05 (1.04 - 1.05) | 1.53 (1.51 - 1.54) | 1.08 (1.07 - 1.09) |
| East | 1.21 (1.19 - 1.22) | 4.79 (4.67 - 4.91) | 0.92 (0.91 - 0.93) | 2.74 (2.71 - 2.78) | 1.29 (1.28 - 1.30) | 1.89 (1.87 - 1.91) | 1.34 (1.33 - 1.35) |
| London | 0.71 (0.69 - 0.73) | 3.08 (2.90 - 3.26) | 0.49 (0.48 - 0.51) | 2.07 (2.01 - 2.14) | 0.71 (0.69 - 0.73) | 1.22 (1.19 - 1.25) | 0.76 (0.74 - 0.77) |
| North East | 1.34 (1.32 - 1.36) | 5.32 (5.13 - 5.51) | 1.06 (1.05 - 1.08) | 2.88 (2.81 - 2.95) | 1.43 (1.41 - 1.45) | 2.09 (2.04 - 2.14) | 1.52 (1.50 - 1.55) |
| North West | 1.39 (1.38 - 1.41) | 5.76 (5.61 - 5.92) | 1.12 (1.11 - 1.14) | 2.81 (2.76 - 2.86) | 1.51 (1.50 - 1.53) | 2.11 (2.08 - 2.14) | 1.53 (1.51 - 1.54) |
| South East | 0.96 (0.93 - 0.98) | 3.73 (3.50 - 3.96) | 0.72 (0.70 - 0.74) | 2.24 (2.17 - 2.30) | 1.00 (0.98 - 1.02) | 1.44 (1.41 - 1.47) | 1.03 (1.02 - 1.05) |
| South West | 1.07 (1.05 - 1.09) | 4.55 (4.37 - 4.74) | 0.81 (0.79 - 0.82) | 2.57 (2.52 - 2.62) | 1.12 (1.11 - 1.13) | 1.61 (1.59 - 1.63) | 1.15 (1.14 - 1.16) |
| West Midlands | 1.32 (1.29 - 1.34) | 5.63 (5.38 - 5.88) | 0.97 (0.95 - 0.99) | 3.00 (2.92 - 3.07) | 1.37 (1.34 - 1.39) | 2.20 (2.15 - 2.26) | 1.49 (1.46 - 1.52) |
| Yorkshire & The Humber | 1.31 (1.30 - 1.33) | 5.46 (5.34 - 5.58) | 1.03 (1.02 - 1.04) | 2.93 (2.89 - 2.97) | 1.41 (1.39 - 1.42) | 2.01 (1.98 - 2.03) | 1.44 (1.43 - 1.45) |
| **IMD quintile** |  |  |  |  |  |  |  |
| 1 (most deprived) | 1.58 (1.57 - 1.59) | 5.27 (5.17 - 5.38) | 1.20 (1.19 - 1.21) | 3.13 (3.10 - 3.17) | 1.66 (1.65 - 1.67) | 2.45 (2.42 - 2.48) | 1.71 (1.69 - 1.72) |
| 2 | 1.33 (1.32 - 1.34) | 5.07 (4.96 - 5.18) | 1.02 (1.01 - 1.03) | 2.84 (2.80 - 2.88) | 1.35 (1.34 - 1.36) | 2.01 (1.99 - 2.03) | 1.38 (1.37 - 1.39) |
| 3 | 1.12 (1.11 - 1.13) | 4.84 (4.72 - 4.95) | 0.87 (0.86 - 0.88) | 2.57 (2.54 - 2.61) | 1.13 (1.12 - 1.14) | 1.65 (1.64 - 1.67) | 1.15 (1.14 - 1.16) |
| 4 | 1.00 (0.99 - 1.01) | 4.65 (4.53 - 4.77) | 0.77 (0.76 - 0.78) | 2.38 (2.34 - 2.41) | 1.02 (1.01 - 1.03) | 1.50 (1.48 - 1.52) | 1.03 (1.02 - 1.04) |
| 5 (least deprived) | 0.86 (0.85 - 0.88) | 4.23 (4.11 - 4.35) | 0.66 (0.65 - 0.67) | 2.18 (2.14 - 2.22) | 0.88 (0.87 - 0.89) | 1.27 (1.26 - 1.29) | 0.90 (0.89 - 0.91) |

The fit note rate was higher in the hospitalised cohorts, and was 6.78 per 100 person-months (95%CI 6.59-6.98) in 2020, 7.19 (95%CI 7.03 - 7.36) in 2021 and 4.13 (95%CI 4.03-4.22) in 2022. In contrast, the fit note rate in the pneumonia cohort was 7.17 (95%CI 7.01-7.34) (**Supplementary Table 3**).

### Fit note rate by demographics

Among people in the COVID-19 cohorts, the first fit note rate was higher in 2020 and lower in 2022 for all demographic groups except people 18-24 years (**Table 2**). Generally, the fit note rate was higher in people aged ≥45 years, women, people of Asian or Asian British ethnicity, and lower in people 18-24 years and living in London. The fit note rate increased with greater deprivation as defined by the IMD quintile. In most cases these patterns reflected those in the general population.

A slightly different pattern was observed for the hospitalised cohorts (**Supplementary Table 3**). People 45-54 years were most likely to receive a fit note, which was older than the pneumonia cohort. Among hospitalised patients the relationship between sex and receiving a fit note differed by year, with women more likely to receive a fit note in 2020 and men more likely in 2021 and 2022. A linear relationship between lesser deprivation and a higher fit note rate was seen for the pneumonia cohort, but not the hospitalised COVID-19 cohorts.

### Cox regression by follow-up period

The overall fit note rate was higher in all COVID-19 cohorts than either their contemporary or 2019 cohorts. Most people in the COVID-19 cohorts who were issued a first fit note received it in the first 30 days post-index date (**Table 3**). The fully adjusted HR representing the average effect over the entire study period was highest comparing the 2020 COVID-19 cohort to the 2020 general population (4.07, 95%CI 4.02-4.12) and lowest comparing the 2022 COVID-19 cohort to the 2022 general population (1.57, 95%CI 1.56-1.58) (**Table 3**). For all comparisons of the COVID-19 population with the general population, the HR was greatest when we restricted the analysis to the first 30 days from the index date, with fully adjusted HRs in this early follow-up period ranging from 5.94 (95%CI 5.85-6.03) comparing the 2020 COVID-19 cohort to the 2020 general population to 2.37 (95%CI 2.34-2.39) comparing the 2022 COVID-19 cohort to the 2022 general population. With longer follow-up periods, the HR attenuated but remained high.

**Table 3.** Crude and adjusted HRs for receipt of first fit note, overall and by follow-up period post-index date.

| Comparison and follow-up period post-index date | Fit note rate per 100 person-months (95%CI) |  | Crude HR (95%CI) | Age-sex adjusted HR (95%CI) | Fully adjusted HR* (95%CI) |
| --- | --- | --- | --- | --- | --- |
|  | COVID-19 cohort | General population |  |  |  |
| **2020 COVID-19 cohort vs 2019 general population** |  |  |  |  |  |
| 0-30 days | 9.35 (9.24 - 9.46) | 1.99 (1.98 - 2.01) | 4.59 (4.52 - 4.65) | 4.77 (4.70 - 4.84) | 4.68 (4.61 - 4.75) |
| 0-90 days | 6.20 (6.14 - 6.27) | 1.48 (1.47 - 1.48) | 3.59 (3.55 - 3.64) | 3.69 (3.64 - 3.73) | 3.63 (3.58 - 3.68) |
| 0-150 days | 5.39 (5.33 - 5.45) | 1.31 (1.30 - 1.32) | 3.35 (3.31 - 3.39) | 3.41 (3.37 - 3.45) | 3.36 (3.32 - 3.40) |
| Overall | 4.88 (4.83 - 4.93) | 1.21 (1.20 - 1.22) | 3.20 (3.16 - 3.24) | 3.23 (3.20 - 3.27) | 3.19 (3.15 - 3.23) |
| **2020 COVID-19 cohort vs 2020 general population** |  |  |  |  |  |
| 0-30 days | 9.35 (9.24 - 9.46) | 1.57 (1.55 - 1.58) | 5.83 (5.74 - 5.92) | 6.14 (6.05 - 6.24) | 5.94 (5.85 - 6.03) |
| 0-90 days | 6.20 (6.14 - 6.27) | 1.12 (1.11 - 1.13) | 4.65 (4.59 - 4.71) | 4.83 (4.77 - 4.89) | 4.67 (4.61 - 4.73) |
| 0-150 days | 5.39 (5.33 - 5.45) | 0.99 (0.99 - 1.00) | 4.35 (4.30 - 4.40) | 4.46 (4.40 - 4.51) | 4.32 (4.27 - 4.38) |
| Overall | 4.88 (4.83 - 4.93) | 0.93 (0.93 - 0.93) | 4.13 (4.08 - 4.18) | 4.19 (4.14 - 4.24) | 4.07 (4.02 - 4.12) |
| **2021 COVID-19 cohort vs 2019 general population** |  |  |  |  |  |
| 0-30 days | 6.51 (6.46 - 6.56) | 1.99 (1.98 - 2.01) | 3.24 (3.21 - 3.28) | 3.33 (3.29 - 3.37) | 3.34 (3.30 - 3.38) |
| 0-90 days | 3.50 (3.48 - 3.52) | 1.48 (1.47 - 1.48) | 2.27 (2.25 - 2.29) | 2.39 (2.37 - 2.41) | 2.38 (2.35 - 2.41) |
| 0-150 days | 2.91 (2.90 - 2.93) | 1.31 (1.30 - 1.32) | 2.06 (2.04 - 2.07) | 2.17 (2.15 - 2.18) | 2.15 (2.13 - 2.17) |
| Overall | 2.66 (2.64 - 2.67) | 1.21 (1.20 - 1.22) | 1.96 (1.95 - 1.98) | 2.04 (2.03 - 2.06) | 2.02 (2.00 - 2.04) |
| **2021 COVID-19 cohort vs 2021 general population** |  |  |  |  |  |
| 0-30 days | 6.51 (6.46 - 6.56) | 1.97 (1.96 - 1.99) | 3.27 (3.24 - 3.31) | 3.43 (3.40 - 3.47) | 3.37 (3.33 - 3.40) |
| 0-90 days | 3.50 (3.48 - 3.52) | 1.47 (1.46 - 1.48) | 2.29 (2.28 - 2.31) | 2.40 (2.38 - 2.42) | 2.35 (2.33 - 2.37) |
| 0-150 days | 2.91 (2.90 - 2.93) | 1.32 (1.31 - 1.33) | 2.07 (2.06 - 2.09) | 2.17 (2.16 - 2.19) | 2.12 (2.10 - 2.13) |
| Overall | 2.66 (2.64 - 2.67) | 1.23 (1.22 - 1.23) | 1.97 (1.96 - 1.99) | 2.06 (2.04 - 2.07) | 1.99 (1.98 - 2.01) |
| **2022 COVID-19 cohort vs 2019 general population** |  |  |  |  |  |
| 0-30 days | 4.87 (4.83 - 4.91) | 1.99 (1.98 - 2.01) | 2.46 (2.43 - 2.48) | 2.14 (2.11 - 2.17) | 2.27 (2.24 - 2.30) |

| Comparison and follow-up period<br>post-index date | Fit note rate per 100 person-months (95%CI) |  | Crude HR (95%CI) | Age-sex adjusted HR<br>(95%CI) | Fully adjusted HR* (95%CI) |
| --- | --- | --- | --- | --- | --- |
|  | COVID-19 cohort | General population |  |  |  |
| 0-90 days | 2.57 (2.55 - 2.58) | 1.48 (1.47 - 1.48) | 1.79 (1.77 - 1.81) | 1.63 (1.62 - 1.65) | 1.73 (1.72 - 1.75) |
| 0-150 days | 2.05 (2.04 - 2.06) | 1.31 (1.30 - 1.32) | 1.65 (1.64 - 1.66) | 1.53 (1.52 - 1.55) | 1.63 (1.61 - 1.64) |
| Overall | 1.73 (1.72 - 1.73) | 1.21 (1.20 - 1.22) | 1.56 (1.55 - 1.57) | 1.46 (1.45 - 1.47) | 1.55 (1.54 - 1.57) |
| **2022 COVID-19 cohort vs 2022**<br>**general population** |  |  |  |  |  |
| 0-30 days | 4.87 (4.83 - 4.91) | 2.09 (2.07 - 2.10) | 2.35 (2.32 - 2.37) | 2.35 (2.33 - 2.38) | 2.37 (2.34 - 2.39) |
| 0-90 days | 2.57 (2.55 - 2.58) | 1.53 (1.52 - 1.54) | 1.73 (1.71 - 1.74) | 1.73 (1.72 - 1.74) | 1.73 (1.73 - 1.76) |
| 0-150 days | 2.05 (2.04 - 2.06) | 1.34 (1.39 - 1.35) | 1.62 (1.60 - 1.63) | 1.62 (1.61 - 1.63) | 1.64 (1.62 - 1.65) |
| Overall | 1.73 (1.72 - 1.73) | 1.23 (1.22 - 1.23) | 1.55 (1.54 - 1.56) | 1.55 (1.54 - 1.56) | 1.57 (1.56 - 1.58) |

^General population age, sex and STP frequency matched.
\*Fully adjusted models include age, sex, IMD quintile, region, ethnicity, obesity, smoking status, hypertension, diabetes, chronic respiratory disease, asthma, chronic cardiac disease, lung cancer, haematological cancer, other cancer, chronic liver disease, other neurological disease, organ transplant, asplenia, HIV, permanent immunodeficiency, and rheumatoid arthritis/systemic lupus erythematosus/psoriasis.

For the hospitalised cohorts, the overall fit note rate was lower in the COVID-19 cohorts than the pneumonia cohort with the highest rates in the first 30 days. The average HR over all follow-up time ranged from 0.62 (95%CI 0.59-0.65) for the 2022 COVID-19 cohort to 0.81 (95%CI 0.77-0.86) for the 2021 cohort (**Supplementary Table 4**). The HR was relatively stable regardless of follow-up period used.

### Cox regression stratified by demographic categories

For all demographics groups, the HR was greatest in 2020 when compared with either a contemporary or 2019 historical comparator (**Figure 1, Supplementary Tables 5-9**). In 2020, large relative increases in fit note rate were observed for people ≥35 years, women, and people of Black or Other ethnicity. The HR was also higher in people in less deprived IMD quintiles; when comparing the 2020 COVID-19 cohort to the 2020 general population, the fully adjusted HR ranged from 3.41 (95%CI 3.33-3.48) in the most deprived quintile to 5.15 (95%CI 4.98-5.32) in the least deprived quintile.

**Figure 1.**
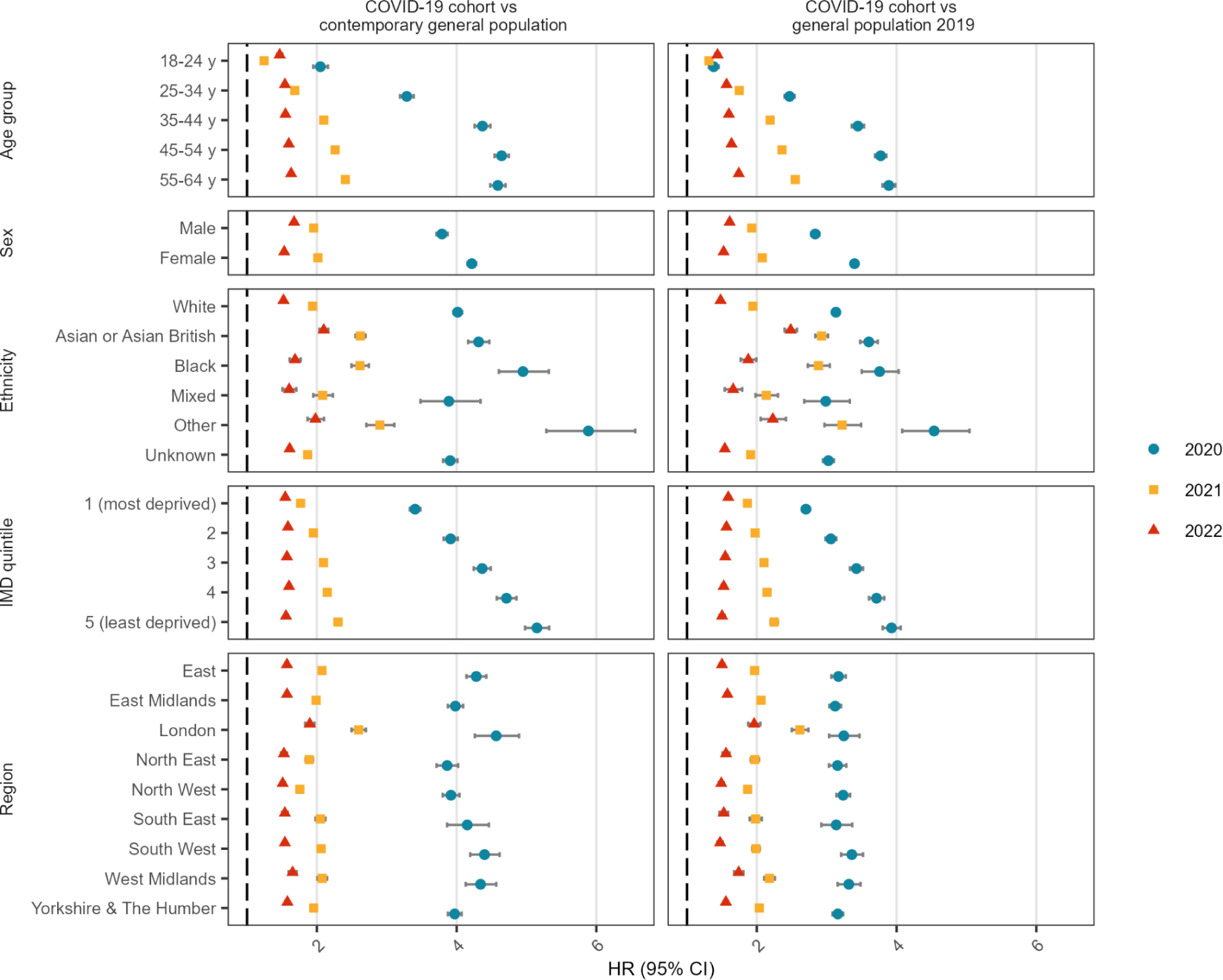
Fully adjusted hazard ratio of first fit note comparing COVID-19 cohorts to their contemporary or historical 2019 general population, stratified by demographic categories. *All models are adjusted for age, sex, ethnicity, IMD quintile and region, but exclude the stratification variable. Models are additionally adjusted for obesity, smoking status, hypertension, diabetes, chronic respiratory disease, asthma, chronic cardiac disease, lung cancer, haematological cancer, other cancer, chronic liver disease, other neurological disease, organ transplant, asplenia, HIV, permanent immunodeficiency, and rheumatoid arthritis/systemic lupus erythematosus/psoriasis

These HRs were lower in 2021 compared with 2020, and more so in 2022. The one exception was people 18-24 years, where the HR compared with the general population was 1.47 (95%CI 1.43-1.51) in 2022, compared with 1.24 (95%CI 1.22-1.27) in 2021. Other patterns that differed in 2022 compared with other years was that no meaningful variation by IMD quintile was observed, and a higher HR was seen in men compared with women. The findings were similar comparing the COVID-19 cohorts with the 2019 comparator.

Among the hospitalised cohorts, the HRs comparing them with the pneumonia cohort were similar in 2020 and 2021, but lower in 2022 (**Supplementary Figure 3, Supplementary Tables 4-8**). A lower fit note rate in the COVID-19 cohorts compared with the pneumonia cohort was observed for most demographic subgroups

## Discussion

### Summary

We found that people with a recorded positive SARS-CoV-2 test or COVID-19 diagnosis had a higher fit note rate than the general population, even after adjusting for demographics and a wide range of clinical characteristics. This increase was greatest in 2020 but continued even into 2022 despite the introduction of vaccines and other effective outpatient COVID-19 treatments. The fit note rate was highest in the first 30 days post-diagnosis, but an increased risk persisted over all follow-up in all years. These findings provide further evidence for the long-term health and economic impact of COVID-19. In contrast, people hospitalised with COVID-19 were less likely to be issued a fit note than people with pneumonia, especially in 2022, suggesting the long-term effects are not worse than comparable serious respiratory infections requiring hospitalisation.

### Strengths and weaknesses

Our data come from a representative sample of 40% of the English population.(13) To better understand fit note patterns, we used COVID-19 cohorts from three different pandemic time periods, with both contemporary and historical comparators, to account for changes in preventive measures (e.g. lockdowns, vaccines) and other disruptions to working patterns (e.g. furlough) that would have impacted on receipt of fit notes over time.

However, there is bias in who gets tested or seeks care for COVID-19 which will have changed as the pandemic evolved,(14) especially after cessation of free testing in April 2022. It is unclear how this would correlate with the likelihood of requesting or requiring a fit note. To mitigate this, we relied on three different methods for identifying COVID-19 cases (positive PCR or LFT test, primary care diagnosis, hospitalisation), but many mild or asymptomatic cases, and cases among people choosing not to get tested or not to record their LFT result will be missed.

We also could not identify whether people were participating in the workforce and included everyone of working age. From 2019 to 2022, the employment rate was 62% in people 18-24 yrs, 85% in people 25-49 years, and 72% in people 50-64 years.(15) It is therefore likely that the true absolute fit note rate will be substantially attenuated, especially among the younger age groups. However, this will only impact on the relative effects if the employment rate differs substantially between the COVID-19 cohorts and general population.

### Findings in context

The pandemic has evolved over time, both in the number of cases, disease severity, and demographic groups most affected. The introduction of vaccination programmes and differences in circulating variants lead to changes in severity and transmissibility of SARS-CoV-2.(16) Additionally, the ending of lockdowns and other preventive measures, cessation of free testing, and differential uptake of vaccines by demographic and clinical subgroups will have impacted on the characteristics of people testing positive for SARS-CoV-2.(17) Consistent with previously reported data,(5,18) we observed that the number of people with evidence of a positive SARS-CoV-2 test was much greater in 2021 and 2022 compared with 2020. Thus, while the fit note rate was nearly twice as high in 2020, the actual number of people issued a fit note was 4.5 times higher in 2022.

In contrast, the fit note rate was lower in the general population in 2020, compared with other years. This is consistent with a previous study of NHS workers which identified a decrease in fit notes for non-COVID related episodes in 2020.(19) There are likely to be several explanations such as changes to working patterns, including furlough and remote working, which would have reduced the need for fit notes. There were also fewer circulating non-COVID-19 respiratory infections in 2020 due to public health measures.(20) It is also important to note that people who were required to isolate due to COVID-19 would be issued an isolation note, rather than a fit note and thus are not counted.

### Policy implications and interpretation

Now several years into the pandemic, vaccines have led to reduced severity of COVID-19, but the number of daily cases remains high,(17) potentially putting many people at risk of long-term symptoms. The incidence of repeat infections is also increasing;(21) while a repeat infection appears to lead to milder acute symptoms,(22,23) other studies have found that the long-term symptom burden increases with the number of reinfections.(24,25) In our study, we did not try and link the reason for the fit note to COVID-19. However, most studies of long-term effects have focussed on surveys and self-report, which can be unrepresentative(26–28) and long COVID is not well coded in electronic health records.(11) Thus, our findings provide a different perspective on the potential long-term consequences of COVID-19 that does not rely on accurate coding of COVID-19 related symptoms.

### Future research

A previous study identified that while the fit note rate decreased overall in 2020, fit notes associated with certain diagnoses, specifically asthma, respiratory conditions, and mental health increased during the COVID-19 pandemic.(19) Similarly, a study of NHS workers found that sickness absences due to psychiatric illness was higher in 2022 compared with pre-pandemic.(29) Further investigation into the duration of fit notes, as well as the diagnoses associated with the issued fit note, whether these changed over time from diagnosis, and how these compare to fit notes issued to people without COVID-19 will help shed light on the most common long-term symptoms experienced.

### Conclusion

Despite undercapture of people with COVID-19 and overestimation of the number of people in the workforce, we have identified a considerable increased risk of fit notes among people with COVID-19 compared with the general population. This extends into an era of high vaccination rates and when the severity of illness from COVID-19 is decreasing. The majority of fit notes were issued within the first 30 days post-diagnosis, suggesting that most COVID-19 related sick leave is associated with the acute phase of the disease. However, a persistent increased risk up to 10 months after the illness demonstrates the ongoing health and economic impact of COVID-19.

## Data Availability

All data were linked, stored and analysed securely using the OpenSAFELY platform, https://www.opensafely.org/, as part of the NHS England OpenSAFELY COVID-19 service. Data include pseudonymised data such as coded diagnoses, medications and physiological parameters. No free text data are included. All code is shared openly for review and re-use under MIT open license (https://github.com/opensafely/long-covid-sick-notes). Detailed pseudonymised patient data are potentially re-identifiable and therefore not shared.

## Ethics approval

This study was approved by the Health Research Authority (Research Ethics Committee reference 20/LO/0651) and by the London School of Hygiene and Tropical Medicine Ethics Board (reference 21863).

## Acknowledgements

We are very grateful for all the support received from the TPP Technical Operations team throughout this work, and for generous assistance from the information governance and database teams at NHS England and the NHS England Transformation Directorate

## Members of OpenSAFELY Collaborative

Sebastian CJ Bacon, Lucy Bridges, Benjamin FC Butler-Cole, Simon Davy, Iain Dillingham, David Evans, Ben Goldacre, Liam Hart, George Hickman, Peter Inglesby, Steven Maude, Jessica Morley, Amir Mehrkar, Thomas O’Dwyer, Rebecca M Smith, Tom Ward, Jon Massey, Milan Wiedemann, Christopher Bates, Jonathan Cockburn, Sam Harper, Frank Hester, John Parry

## Members of National Core Studies Collaborative

Nishi Chaturvedi, Chloe Park, Alisia Carnemolla, Dylan Williams, Anika Knueppel, Andy Boyd, Emma L Turner, Katharine M Evans, Richard Thomas, Samantha Berman, Stela McLachlan, Matthew Crane, Rebecca Whitehorn, Jacqui Oakley, Diane Foster, Hannah Woodward, Kirsteen C Campbell, Nicholas Timpson, Alex Kwong, Ana Goncalves Soares, Gareth Griffith, Renin Toms, Louise Jones, Annie Herbert, Ruth Mitchell, Tom Palmer, Jonathan Sterne, Venexia Walker, Lizzie Huntley, Laura Fox, Rachel Denholm, Rochelle Knight, Kate, Northstone, Arun, Kanagaratnam, Elsie Horne, Harriet Forbes, Teri North, Kurt, Taylor, Marwa AL Arab, Scott Walker, Jose IC Coronado, Arun S Karthikeyan, George, Ploubidis, Bettina Moltrecht, Charlotte Booth, Sam Parsons, Bozena Wielgoszewska, Charis Bridger-Staatz, Claire Steves, Ellen Thompson, Paz Garcia, Nathan Cheetham, Ruth Bowyer, Maxim Freydin, Amy Roberts, Ben Goldacre, Alex Walker, Jess Morley, William Hulme, Linda Nab, Louis Fisher, Brian MacKenna, Colm Andrews, Helen Curtis, Lisa Hopcroft, Amelia Green, Praveetha Patalay, Jane Maddock, Kishan Patel, Jean Stafford, Wels Jacques, Kate Tilling, John Macleod, Eoin McElroy, Anoop Shah Richard Silverwood, Spiros Denaxas, Robin Flaig, Daniel McCartney, Archie Campbell, Laurie Tomlinson, John Tazare, Bang Zheng, Liam Smeeth, Emily Herrett, Thomas Cowling, Kate Mansfield, Ruth E Costello, Kevin Wang, Kathryn Mansfield, Viyaasan Mahalingasivam, Ian Douglas, Sinead Langan, Sinead Brophy, Michael Parker, Jonathan Kennedy, Rosie McEachan, John Wright, Kathryn Willan, Ellena Badrick, Gillian Santorelli, Tiffany Yang, Bo Hou, Andrew Steptoe, Giorgio Di Gessa, Jingmin Zhu, Paola Zaninotto, Angela Wood, Genevieve Cezard, Samantha Ip, Tom Bolton, Alexia Sampri, Elena Rafeti, Fatima Almaghrabi, Aziz Sheikh, Syed A Shah, Vittal Katikireddi, Richard Shaw, Olivia Hamilton, Michael Green, Theocharis Kromydas, Daniel Kopasker, Felix Greaves, Robert Willans, Fiona Glen, Steve Sharp, Alun Hughes, Andrew Wong, Lee Hamill Howes, Alicja Rapala, Lidia Nigrelli, Fintan McArdle, Chelsea Beckford, Betty Raman, Richard Dobson, Amos Folarin, Callum Stewart, Yatharth Ranjan, Jd Carpentieri, Laura Sheard, Chao Fang, Sarah Baz, Andy Gibson, John Kellas, Stefan Neubauer, Stefan Piechnik, Elena Lukaschuk, Laura C Saunders, James M Wild, Stephen Smith, Peter Jezzard, Elizabeth Tunnicliffe, Zeena-Britt Sanders, Lucy Finnigan, Vanessa Ferreira, Mark Green, Rebecca Rhead, Milla Kibble, Yinghui Wei, Agnieszka Lemanska, Francisco Perez-Reche, Dominik Piehlmaier, Lucy Teece, Edward Parker

## Administrative

### Funding

The OpenSAFELY Platform is supported by grants from the Wellcome Trust (222097/Z/20/Z) and MRC (MR/V015757/1, MC_PC-20059, MR/W016729/1). In addition, development of OpenSAFELY has been funded by the Longitudinal Health and Wellbeing strand of the National Core Studies programme (MC_PC_20030: MC_PC_20059), The NIHR funded CONVALESCENCE programme (COV-LT-0009), NIHR (NIHR135559, COV-LT2-0073), and the Data and Connectivity National Core Study funded by UK Research and Innovation (MC_PC_20058), and Health Data Research UK (HDRUK2021.000, 2021.0157).

The views expressed are those of the authors and not necessarily those of the NIHR, NHS England, Public Health England or the Department of Health and Social Care. Funders had no role in the study design, collection, analysis, and interpretation of data; in the writing of the report; and in the decision to submit the article for publication.

### Information governance and ethical approval

NHS England is the data controller of the NHS England OpenSAFELY COVID-19 Service; TPP is the data processor; all study authors using OpenSAFELY have the approval of NHS England.(1) This implementation of OpenSAFELY is hosted within the TPP environment which is accredited to the ISO 27001 information security standard and is NHS IG Toolkit compliant.(2)

Patient data has been pseudonymised for analysis and linkage using industry standard cryptographic hashing techniques; all pseudonymised datasets transmitted for linkage onto OpenSAFELY are encrypted; access to the NHS England OpenSAFELY COVID-19 service is via a virtual private network (VPN) connection; the researchers hold contracts with NHS England and only access the platform to initiate database queries and statistical models; all database activity is logged; only aggregate statistical outputs leave the platform environment following best practice for anonymisation of results such as statistical disclosure control for low cell counts.(3)

The service adheres to the obligations of the UK General Data Protection Regulation (UK GDPR) and the Data Protection Act 2018. The service previously operated under notices initially issued in February 2020 by the the Secretary of State under Regulation 3(4) of the Health Service (Control of Patient Information) Regulations 2002 (COPI Regulations), which required organisations to process confidential patient information for COVID-19 purposes; this set aside the requirement for patient consent.(4) As of 1 July 2023, the Secretary of State has requested that NHS England continue to operate the Service under the COVID-19 Directions 2020.(5) In some cases of data sharing, the common law duty of confidence is met using, for example, patient consent or support from the Health Research Authority Confidentiality Advisory Group.(6)

Taken together, these provide the legal bases to link patient datasets using the service. GP practices, which provide access to the primary care data, are required to share relevant health information to support the public health response to the pandemic, and have been informed of how the service operates.

1. NHS Digital. The NHS England OpenSAFELY COVID-19 service - privacy notice [Internet]. 2023 [cited 2023 Jul 5]. Available from: https://digital.nhs.uk/coronavirus/coronavirus-covid-19-response-information-governance-hub/the-nhs-england-opensafely-covid-19-service-privacy-notice
2. NHS Digital. NHS Digital. 2023 [cited 2023 Jul 5]. Data Security and Protection Toolkit. Available from: https://digital.nhs.uk/data-and-information/looking-after-information/data-security-and-information-governance/data-security-and-protection-toolkit
3. NHS Digital [Internet]. [cited 2023 Mar 6]. ISB1523: Anonymisation Standard for Publishing Health and Social Care Data. Available from: https://digital.nhs.uk/data-and-information/information-standards/information-standards-and-data-collections-including-extractions/publications-and-notifications/standards-and-collections/isb1523-anonymisation-standard-for-publishing-health-and-social-care-data
4. UK Department of Health and Social Care. GOV.UK. 2022 [cited 2023 Jul 5]. [Withdrawn] Coronavirus (COVID-19): notice under regulation 3(4) of the Health Service (Control of Patient Information) Regulations 2002 – general. Available from: https://www.gov.uk/government/publications/coronavirus-covid-19-notification-of-data-controllers-to-share-information/coronavirus-covid-19-notice-under-regulation-34-of-the-health-service-control-of-patient-information-regulations-2002-general--2
5. NHS Digital. NHS Digital. 2022 [cited 2023 Jul 5]. Secretary of State for Health and Social Care: COVID-19 Public Health Directions 2020. Available from: https://digital.nhs.uk/about-nhs-digital/corporate-information-and-documents/directions-and-data-provision-notices/secretary-of-state-directions/covid-19-public-health-directions-2020
6. NHS Health Research Authority. Health Research Authority. [cited 2023 Jul 5]. Confidentiality Advisory Group. Available from: https://www.hra.nhs.uk/about-us/committees-and-services/confidentiality-advisory-group/

### Data access and verification

Access to the underlying identifiable and potentially re-identifiable pseudonymised electronic health record data is tightly governed by various legislative and regulatory frameworks, and restricted by best practice. The data in OpenSAFELY is drawn from General Practice data across England where TPP is the Data Processor. TPP developers (CB, JC, JP, FH, and SH) initiate an automated process to create pseudonymised records in the core OpenSAFELY database, which are copies of key structured data tables in the identifiable records. These are linked onto key external data resources that have also been pseudonymised via SHA-512 one-way hashing of NHS numbers using a shared salt. DataLab developers and PIs holding contracts with NHS England have access to the OpenSAFELY pseudonymised data tables as needed to develop the OpenSAFELY tools. These tools in turn enable researchers with OpenSAFELY Data Access Agreements to write and execute code for data management and data analysis without direct access to the underlying raw pseudonymised patient data, and to review the outputs of this code. All code for the full data management pipeline—from raw data to completed results for this analysis—and for the OpenSAFELY platform as a whole is available for review at github.com/OpenSAFELY.

### Author contributions

Conceptualization: AJW, LAT, JM, FG, SD; Data curation: ALS, RYP, AJW, SCJB, ID, CB, BG; Formal analysis: ALS, RYP, AJW, JT; Funding acquisition: BG; Investigation: ALS, AJW, RYP; Methodology: ALS, AJW, LAT, RYP, JM, FG, JT, KB; Project administration: AJW, LAT, AM, BG; Resources: AJW, ID, SCJB, BG; Software: ALS, RYP, AJW, JT, SCJB, ID, CB; Supervision: AJW, LAT, BMK, AM, BG; Validation: ALS, RYP, AJW; Visualisation: ALS, RYP, AJW; Writing - original draft: ALS, RYP, AJW; Writing - review & editing: All authors. All authors gave final approval of the version to be published and agree to be accountable for all aspects of the work. All authors confirm that they had full access to all the data in the study and accept responsibility to submit for publication.

## Supplementary Files

**Supplementary Figure 1.**
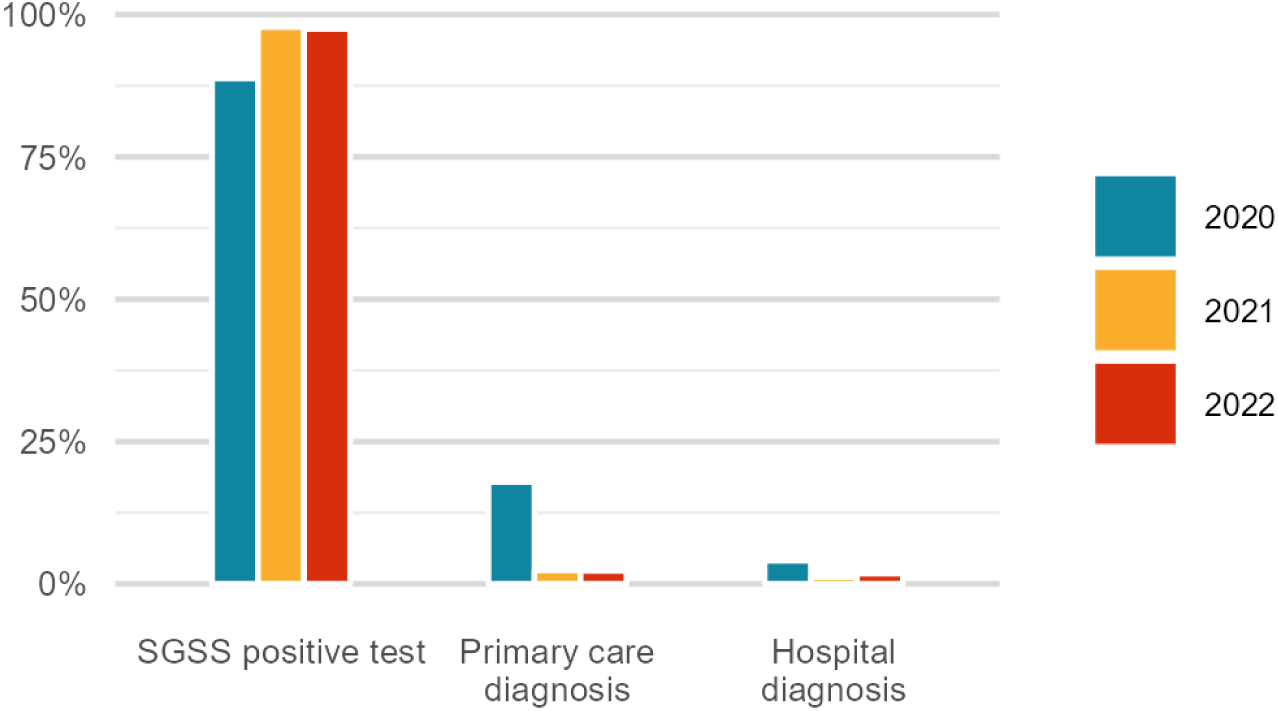
Source of first recorded positive SARS-CoV-2 test or COVID-19 diagnosis, by year. Some people have been identified by multiple sources and therefore the percentages within years may not add up to 100%. SGSS = Second Generation Surveillance System.

**Supplementary Figure 2.**
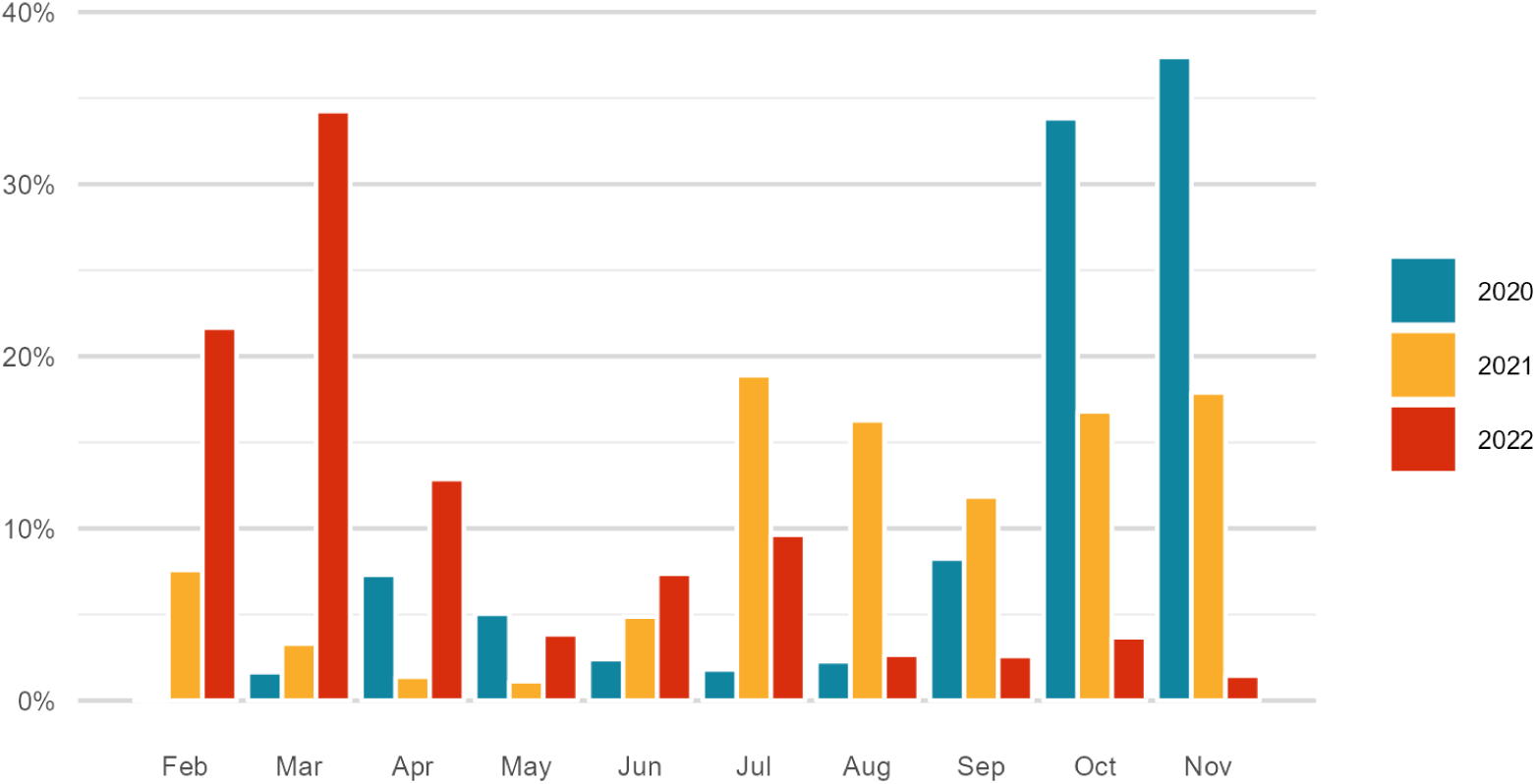
Month of first recorded positive SARS-CoV-2 test or COVID-19 diagnosis by year

**Supplementary Table 1.** Demographic and clinical characteristics of hospitalised cohorts, 1 Feb - 30 Nov of each year. All counts rounded to nearest 7.

|  | COVID-19 hospitalised cohorts |  |  | Pneumonia hospitalised cohort |
| --- | --- | --- | --- | --- |
|  | 2020 | 2021 | 2022 | 2019 |
|  | n (%) | n (%) | n (%) | n (%) |
| **Total** | 22,015 (100.0) | 30,205 (100.0) | 34,692 (100.0) | 29,673 (100.0) |
| **Age** |  |  |  |  |
| 18-24 y | 819 (3.7) | 2107 (7.0) | 2485 (7.2) | 1106 (3.7) |
| 25-34 y | 2429 (11.0) | 5635 (18.7) | 6419 (18.5) | 2618 (8.8) |
| 35-44 y | 3465 (15.7) | 6307 (20.9) | 6006 (17.3) | 4158 (14.0) |
| 45-54 y | 6328 (28.7) | 7329 (24.3) | 7623 (22.0) | 7798 (26.3) |
| 55-64 y | 8974 (40.8) | 8827 (29.2) | 12,166 (35.1) | 13,986 (47.1) |
| **Sex** |  |  |  |  |
| Female | 10,059 (45.7) | 15,442 (51.1) | 20,531 (59.2) | 14,294 (48.2) |
| Male | 11,956 (54.3) | 14,763 (48.9) | 14,161 (40.8) | 15,379 (51.8) |
| **Ethnicity** |  |  |  |  |
| White | 12,376 (56.2) | 18,305 (60.6) | 24,542 (70.7) | 19,817 (66.8) |
| Asian or Asian British | 3395 (15.4) | 3374 (11.2) | 2289 (6.6) | 1589 (5.4) |
| Black | 1008 (4.6) | 1260 (4.2) | 805 (2.3) | 553 (1.9) |
| Mixed | 343 (1.6) | 532 (1.8) | 406 (1.2) | 217 (0.7) |
| Other | 532 (2.4) | 623 (2.1) | 511 (1.5) | 273 (0.9) |
| Unknown | 4361 (19.8) | 6118 (20.3) | 6146 (17.7) | 7217 (24.3) |
| **IMD** |  |  |  |  |
| 1 (most deprived) | 7077 (32.1) | 10,199 (33.8) | 9478 (27.3) | 8974 (30.2) |
| 2 | 5040 (22.9) | 6839 (22.6) | 7525 (21.7) | 6636 (22.4) |
| 3 | 3962 (18.0) | 5446 (18.0) | 6951 (20.0) | 5775 (19.5) |
| 4 | 3269 (14.8) | 4319 (14.3) | 5957 (17.2) | 4662 (15.7) |
| 5 (least deprived) | 2667 (12.1) | 3395 (11.2) | 4774 (13.8) | 3626 (12.2) |
| **Region** |  |  |  |  |
| East | 3885 (17.6) | 5341 (17.7) | 7238 (20.9) | 5866 (19.8) |
| East Midlands | 4375 (19.9) | 6090 (20.2) | 6776 (19.5) | 5698 (19.2) |
| London | 1680 (7.6) | 1715 (5.7) | 1848 (5.3) | 1624 (5.5) |
| North East | 1568 (7.1) | 2044 (6.8) | 2219 (6.4) | 1715 (5.8) |
| North West | 2457 (11.2) | 3339 (11.1) | 3542 (10.2) | 2926 (9.9) |
| South East | 854 (3.9) | 1421 (4.7) | 2359 (6.8) | 1701 (5.7) |
| South West | 1631 (7.4) | 3080 (10.2) | 4326 (12.5) | 3857 (13.0) |
| West Midlands | 1547 (7.0) | 2037 (6.7) | 1771 (5.1) | 1463 (4.9) |
| Yorkshire & The Humber | 4025 (18.3) | 5145 (17.0) | 4613 (13.3) | 4816 (16.2) |
| **Health conditions** |  |  |  |  |
| Obesity | 9870 (44.8) | 12,600 (41.7) | 11,452 (33.0) | 9877 (33.3) |
| Hypertension | 6594 (30.0) | 6552 (21.7) | 8435 (24.3) | 8386 (28.3) |
| Diabetes | 5726 (26.0) | 5817 (19.3) | 6846 (19.7) | 6146 (20.7) |
| Asthma | 4907 (22.3) | 6755 (22.4) | 8694 (25.1) | 7581 (25.5) |
| Other chronic respiratory disease | 1645 (7.5) | 1722 (5.7) | 3304 (9.5) | 5460 (18.4) |
| Chronic cardiac disease | 2324 (10.6) | 2198 (7.3) | 3780 (10.9) | 4095 (13.8) |
| Lung cancer | 154 (0.7) | 126 (0.4) | 301 (0.9) | 735 (2.5) |
| Haematological cancer | 371 (1.7) | 469 (1.6) | 1162 (3.3) | 882 (3.0) |
| Other cancer | 1323 (6.0) | 1358 (4.5) | 3199 (9.2) | 3325 (11.2) |
| Chronic liver disease | 623 (2.8) | 595 (2.0) | 1582 (4.6) | 1477 (5.0) |
| Other neurological disease | 651 (3.0) | 595 (2.0) | 1841 (5.3) | 1260 (4.2) |
| Organ transplant | 280 (1.3) | 455 (1.5) | 1253 (3.6) | 490 (1.7) |
| Asplenia | 77 (0.3) | 91 (0.3) | 203 (0.6) | 189 (0.6) |
| HIV | 14 (0.1) | 14 (0.0) | 49 (0.1) | 21 (0.1) |
| Other permanent immunodeficiency | 49 (0.2) | 70 (0.2) | 196 (0.6) | 91 (0.3) |
| Rheumatoid arthritis / SLE / Psoriasis | 1505 (6.8) | 1967 (6.5) | 3066 (8.8) | 2569 (8.7) |
| **Smoking status** |  |  |  |  |
| Current | 3171 (14.4) | 4788 (15.9) | 9093 (26.2) | 11,039 (37.2) |
| Former | 7791 (35.4) | 10,654 (35.3) | 11,277 (32.5) | 9233 (31.1) |
| Never | 10,752 (48.8) | 14,021 (46.4) | 13,636 (39.3) | 9058 (30.5) |
| Missing | 301 (1.4) | 742 (2.5) | 679 (2.0) | 336 (1.1) |

^age, sex and STP frequency matched; HIV = human immunodeficiency virus; SLE = systemic lupus erythematosus

**Supplementary Table 2.**
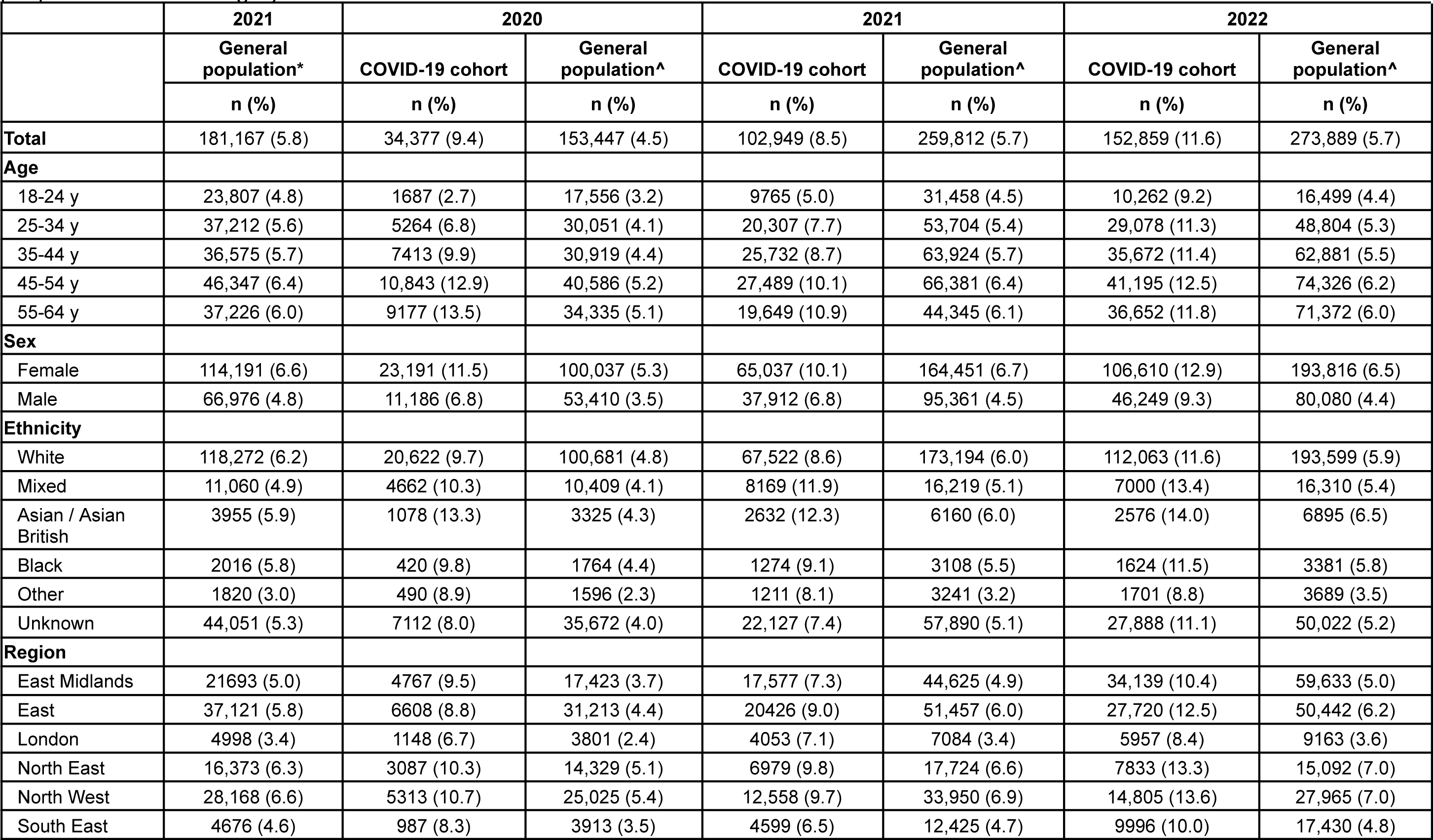
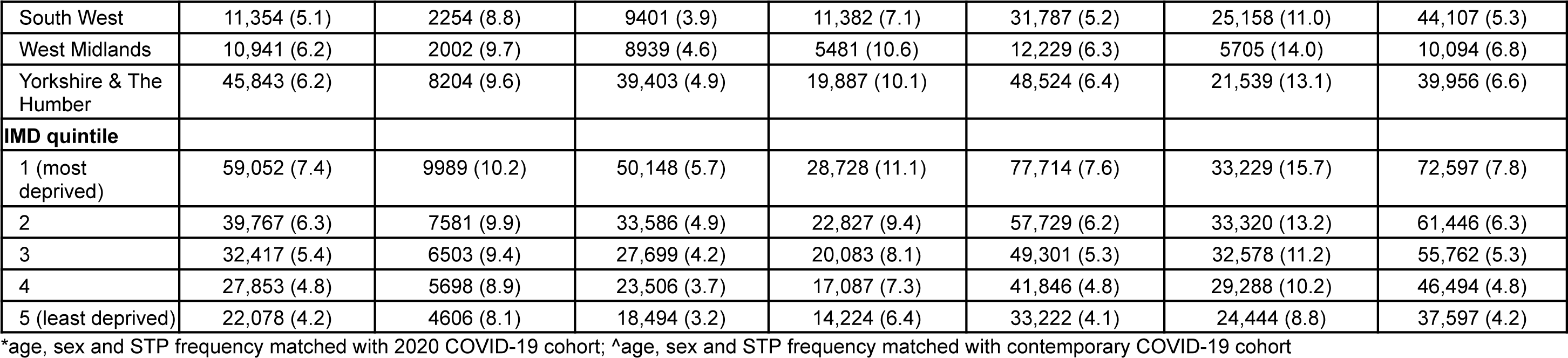
Number and percentage of people with a fit note in each cohort by demographic categories. The denominator is all people within each category. All counts rounded to the nearest 7.

|  | 2021 | 2020 |  | 2021 |  | 2022 |  |
| --- | --- | --- | --- | --- | --- | --- | --- |
|  | General population* | COVID-19 cohort | General population^ | COVID-19 cohort | General population^ | COVID-19 cohort | General population^ |
|  | n (%) | n (%) | n (%) | n (%) | n (%) | n (%) | n (%) |
| **Total** | 181,167 (5.8) | 34,377 (9.4) | 153,447 (4.5) | 102,949 (8.5) | 259,812 (5.7) | 152,859 (11.6) | 273,889 (5.7) |
| **Age** |  |  |  |  |  |  |  |
| 18-24 y | 23,807 (4.8) | 1687 (2.7) | 17,556 (3.2) | 9765 (5.0) | 31,458 (4.5) | 10,262 (9.2) | 16,499 (4.4) |
| 25-34 y | 37,212 (5.6) | 5264 (6.8) | 30,051 (4.1) | 20,307 (7.7) | 53,704 (5.4) | 29,078 (11.3) | 48,804 (5.3) |
| 35-44 y | 36,575 (5.7) | 7413 (9.9) | 30,919 (4.4) | 25,732 (8.7) | 63,924 (5.7) | 35,672 (11.4) | 62,881 (5.5) |
| 45-54 y | 46,347 (6.4) | 10,843 (12.9) | 40,586 (5.2) | 27,489 (10.1) | 66,381 (6.4) | 41,195 (12.5) | 74,326 (6.2) |
| 55-64 y | 37,226 (6.0) | 9177 (13.5) | 34,335 (5.1) | 19,649 (10.9) | 44,345 (6.1) | 36,652 (11.8) | 71,372 (6.0) |
| **Sex** |  |  |  |  |  |  |  |
| Female | 114,191 (6.6) | 23,191 (11.5) | 100,037 (5.3) | 65,037 (10.1) | 164,451 (6.7) | 106,610 (12.9) | 193,816 (6.5) |
| Male | 66,976 (4.8) | 11,186 (6.8) | 53,410 (3.5) | 37,912 (6.8) | 95,361 (4.5) | 46,249 (9.3) | 80,080 (4.4) |
| **Ethnicity** |  |  |  |  |  |  |  |
| White | 118,272 (6.2) | 20,622 (9.7) | 100,681 (4.8) | 67,522 (8.6) | 173,194 (6.0) | 112,063 (11.6) | 193,599 (5.9) |
| Mixed | 11,060 (4.9) | 4662 (10.3) | 10,409 (4.1) | 8169 (11.9) | 16,219 (5.1) | 7000 (13.4) | 16,310 (5.4) |
| Asian / Asian British | 3955 (5.9) | 1078 (13.3) | 3325 (4.3) | 2632 (12.3) | 6160 (6.0) | 2576 (14.0) | 6895 (6.5) |
| Black | 2016 (5.8) | 420 (9.8) | 1764 (4.4) | 1274 (9.1) | 3108 (5.5) | 1624 (11.5) | 3381 (5.8) |
| Other | 1820 (3.0) | 490 (8.9) | 1596 (2.3) | 1211 (8.1) | 3241 (3.2) | 1701 (8.8) | 3689 (3.5) |
| Unknown | 44,051 (5.3) | 7112 (8.0) | 35,672 (4.0) | 22,127 (7.4) | 57,890 (5.1) | 27,888 (11.1) | 50,022 (5.2) |
| **Region** |  |  |  |  |  |  |  |
| East Midlands | 21693 (5.0) | 4767 (9.5) | 17,423 (3.7) | 17,577 (7.3) | 44,625 (4.9) | 34,139 (10.4) | 59,633 (5.0) |
| East | 37,121 (5.8) | 6608 (8.8) | 31,213 (4.4) | 20426 (9.0) | 51,457 (6.0) | 27,720 (12.5) | 50,442 (6.2) |
| London | 4998 (3.4) | 1148 (6.7) | 3801 (2.4) | 4053 (7.1) | 7084 (3.4) | 5957 (8.4) | 9163 (3.6) |
| North East | 16,373 (6.3) | 3087 (10.3) | 14,329 (5.1) | 6979 (9.8) | 17,724 (6.6) | 7833 (13.3) | 15,092 (7.0) |
| North West | 28,168 (6.6) | 5313 (10.7) | 25,025 (5.4) | 12,558 (9.7) | 33,950 (6.9) | 14,805 (13.6) | 27,965 (7.0) |
| South East | 4676 (4.6) | 987 (8.3) | 3913 (3.5) | 4599 (6.5) | 12,425 (4.7) | 9996 (10.0) | 17,430 (4.8) |
| South West | 11,354 (5.1) | 2254 (8.8) | 9401 (3.9) | 11,382 (7.1) | 31,787 (5.2) | 25,158 (11.0) | 44,107 (5.3) |
| West Midlands | 10,941 (6.2) | 2002 (9.7) | 8939 (4.6) | 5481 (10.6) | 12,229 (6.3) | 5705 (14.0) | 10,094 (6.8) |
| Yorkshire & The Humber | 45,843 (6.2) | 8204 (9.6) | 39,403 (4.9) | 19,887 (10.1) | 48,524 (6.4) | 21,539 (13.1) | 39,956 (6.6) |
| **IMD quintile** |  |  |  |  |  |  |  |
| 1 (most deprived) | 59,052 (7.4) | 9989 (10.2) | 50,148 (5.7) | 28,728 (11.1) | 77,714 (7.6) | 33,229 (15.7) | 72,597 (7.8) |
| 2 | 39,767 (6.3) | 7581 (9.9) | 33,586 (4.9) | 22,827 (9.4) | 57,729 (6.2) | 33,320 (13.2) | 61,446 (6.3) |
| 3 | 32,417 (5.4) | 6503 (9.4) | 27,699 (4.2) | 20,083 (8.1) | 49,301 (5.3) | 32,578 (11.2) | 55,762 (5.3) |
| 4 | 27,853 (4.8) | 5698 (8.9) | 23,506 (3.7) | 17,087 (7.3) | 41,846 (4.8) | 29,288 (10.2) | 46,494 (4.8) |
| 5 (least deprived) | 22,078 (4.2) | 4606 (8.1) | 18,494 (3.2) | 14,224 (6.4) | 33,222 (4.1) | 24,444 (8.8) | 37,597 (4.2) |

\*age, sex and STP frequency matched with 2020 COVID-19 cohort; ^age, sex and STP frequency matched with contemporary COVID-19 cohort

**Supplementary Table 3.**
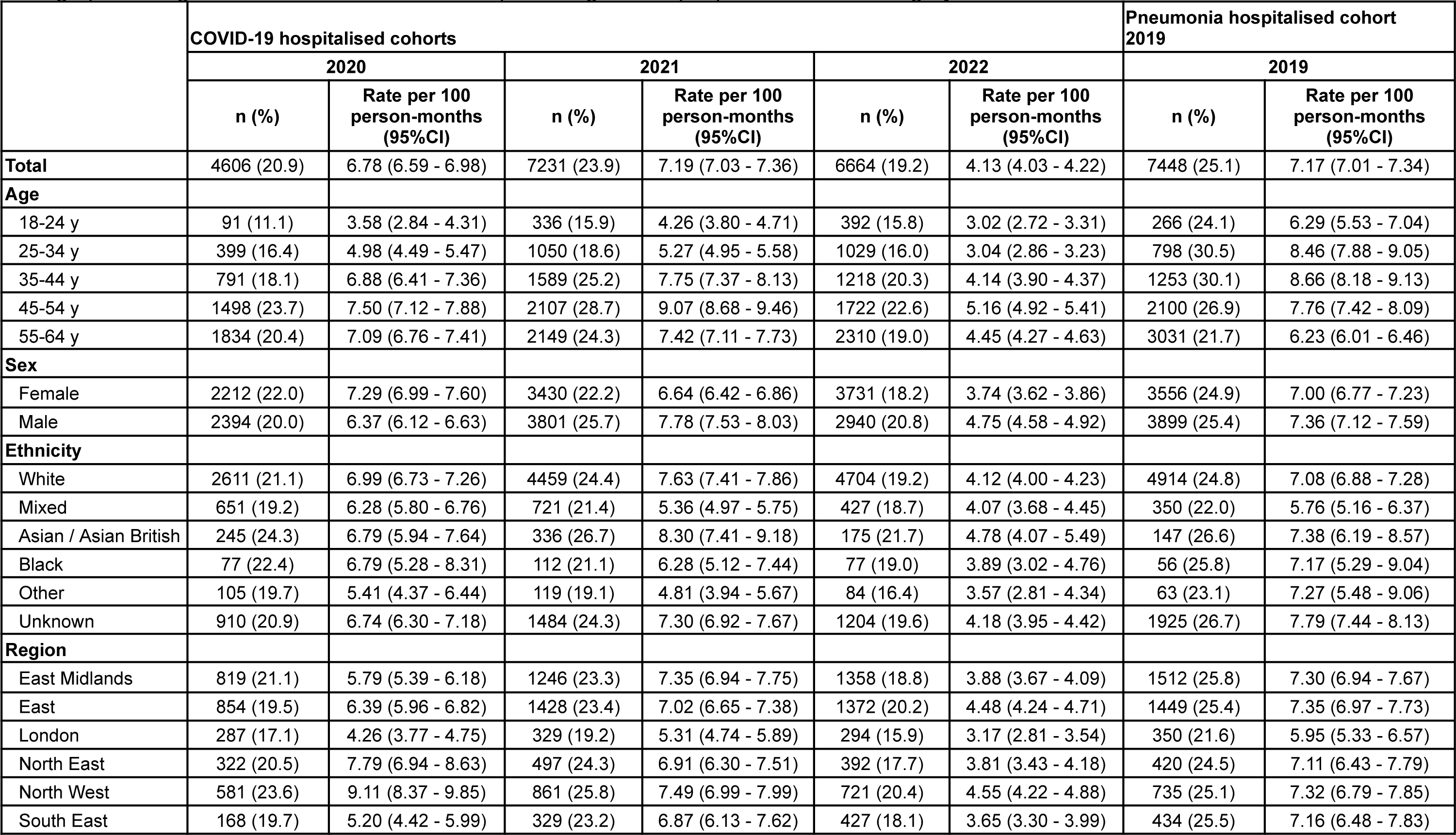
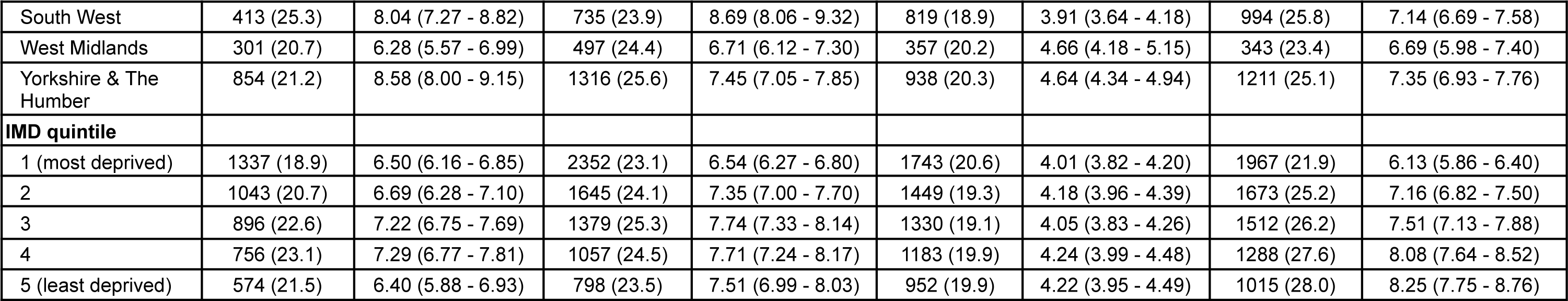
Number of people issued a fit note and rate of first fit note per 100 person-months for hospitalised cohorts by demographic categories. The denominator for the percentages is all people within each category. All counts rounded to the nearest 7.

|  | COVID-19 hospitalised cohorts |  |  |  |  |  | Pneumonia hospitalised cohort 2019 |  |
| --- | --- | --- | --- | --- | --- | --- | --- | --- |
|  | 2020 |  | 2021 |  | 2022 |  | 2019 |  |
|  | n (%) | Rate per 100 person-months (95%CI) | n (%) | Rate per 100 person-months (95%CI) | n (%) | Rate per 100 person-months (95%CI) | n (%) | Rate per 100 person-months (95%CI) |
| **Total** | 4606 (20.9) | 6.78 (6.59 - 6.98) | 7231 (23.9) | 7.19 (7.03 - 7.36) | 6664 (19.2) | 4.13 (4.03 - 4.22) | 7448 (25.1) | 7.17 (7.01 - 7.34) |
| **Age** |  |  |  |  |  |  |  |  |
| 18-24 y | 91 (11.1) | 3.58 (2.84 - 4.31) | 336 (15.9) | 4.26 (3.80 - 4.71) | 392 (15.8) | 3.02 (2.72 - 3.31) | 266 (24.1) | 6.29 (5.53 - 7.04) |
| 25-34 y | 399 (16.4) | 4.98 (4.49 - 5.47) | 1050 (18.6) | 5.27 (4.95 - 5.58) | 1029 (16.0) | 3.04 (2.86 - 3.23) | 798 (30.5) | 8.46 (7.88 - 9.05) |
| 35-44 y | 791 (18.1) | 6.88 (6.41 - 7.36) | 1589 (25.2) | 7.75 (7.37 - 8.13) | 1218 (20.3) | 4.14 (3.90 - 4.37) | 1253 (30.1) | 8.66 (8.18 - 9.13) |
| 45-54 y | 1498 (23.7) | 7.50 (7.12 - 7.88) | 2107 (28.7) | 9.07 (8.68 - 9.46) | 1722 (22.6) | 5.16 (4.92 - 5.41) | 2100 (26.9) | 7.76 (7.42 - 8.09) |
| 55-64 y | 1834 (20.4) | 7.09 (6.76 - 7.41) | 2149 (24.3) | 7.42 (7.11 - 7.73) | 2310 (19.0) | 4.45 (4.27 - 4.63) | 3031 (21.7) | 6.23 (6.01 - 6.46) |
| **Sex** |  |  |  |  |  |  |  |  |
| Female | 2212 (22.0) | 7.29 (6.99 - 7.60) | 3430 (22.2) | 6.64 (6.42 - 6.86) | 3731 (18.2) | 3.74 (3.62 - 3.86) | 3556 (24.9) | 7.00 (6.77 - 7.23) |
| Male | 2394 (20.0) | 6.37 (6.12 - 6.63) | 3801 (25.7) | 7.78 (7.53 - 8.03) | 2940 (20.8) | 4.75 (4.58 - 4.92) | 3899 (25.4) | 7.36 (7.12 - 7.59) |
| **Ethnicity** |  |  |  |  |  |  |  |  |
| White | 2611 (21.1) | 6.99 (6.73 - 7.26) | 4459 (24.4) | 7.63 (7.41 - 7.86) | 4704 (19.2) | 4.12 (4.00 - 4.23) | 4914 (24.8) | 7.08 (6.88 - 7.28) |
| Mixed | 651 (19.2) | 6.28 (5.80 - 6.76) | 721 (21.4) | 5.36 (4.97 - 5.75) | 427 (18.7) | 4.07 (3.68 - 4.45) | 350 (22.0) | 5.76 (5.16 - 6.37) |
| Asian / Asian British | 245 (24.3) | 6.79 (5.94 - 7.64) | 336 (26.7) | 8.30 (7.41 - 9.18) | 175 (21.7) | 4.78 (4.07 - 5.49) | 147 (26.6) | 7.38 (6.19 - 8.57) |
| Black | 77 (22.4) | 6.79 (5.28 - 8.31) | 112 (21.1) | 6.28 (5.12 - 7.44) | 77 (19.0) | 3.89 (3.02 - 4.76) | 56 (25.8) | 7.17 (5.29 - 9.04) |
| Other | 105 (19.7) | 5.41 (4.37 - 6.44) | 119 (19.1) | 4.81 (3.94 - 5.67) | 84 (16.4) | 3.57 (2.81 - 4.34) | 63 (23.1) | 7.27 (5.48 - 9.06) |
| Unknown | 910 (20.9) | 6.74 (6.30 - 7.18) | 1484 (24.3) | 7.30 (6.92 - 7.67) | 1204 (19.6) | 4.18 (3.95 - 4.42) | 1925 (26.7) | 7.79 (7.44 - 8.13) |
| **Region** |  |  |  |  |  |  |  |  |
| East Midlands | 819 (21.1) | 5.79 (5.39 - 6.18) | 1246 (23.3) | 7.35 (6.94 - 7.75) | 1358 (18.8) | 3.88 (3.67 - 4.09) | 1512 (25.8) | 7.30 (6.94 - 7.67) |
| East | 854 (19.5) | 6.39 (5.96 - 6.82) | 1428 (23.4) | 7.02 (6.65 - 7.38) | 1372 (20.2) | 4.48 (4.24 - 4.71) | 1449 (25.4) | 7.35 (6.97 - 7.73) |
| London | 287 (17.1) | 4.26 (3.77 - 4.75) | 329 (19.2) | 5.31 (4.74 - 5.89) | 294 (15.9) | 3.17 (2.81 - 3.54) | 350 (21.6) | 5.95 (5.33 - 6.57) |
| North East | 322 (20.5) | 7.79 (6.94 - 8.63) | 497 (24.3) | 6.91 (6.30 - 7.51) | 392 (17.7) | 3.81 (3.43 - 4.18) | 420 (24.5) | 7.11 (6.43 - 7.79) |
| North West | 581 (23.6) | 9.11 (8.37 - 9.85) | 861 (25.8) | 7.49 (6.99 - 7.99) | 721 (20.4) | 4.55 (4.22 - 4.88) | 735 (25.1) | 7.32 (6.79 - 7.85) |
| South East | 168 (19.7) | 5.20 (4.42 - 5.99) | 329 (23.2) | 6.87 (6.13 - 7.62) | 427 (18.1) | 3.65 (3.30 - 3.99) | 434 (25.5) | 7.16 (6.48 - 7.83) |
| South West | 413 (25.3) | 8.04 (7.27 - 8.82) | 735 (23.9) | 8.69 (8.06 - 9.32) | 819 (18.9) | 3.91 (3.64 - 4.18) | 994 (25.8) | 7.14 (6.69 - 7.58) |
| West Midlands | 301 (20.7) | 6.28 (5.57 - 6.99) | 497 (24.4) | 6.71 (6.12 - 7.30) | 357 (20.2) | 4.66 (4.18 - 5.15) | 343 (23.4) | 6.69 (5.98 - 7.40) |
| Yorkshire & The Humber | 854 (21.2) | 8.58 (8.00 - 9.15) | 1316 (25.6) | 7.45 (7.05 - 7.85) | 938 (20.3) | 4.64 (4.34 - 4.94) | 1211 (25.1) | 7.35 (6.93 - 7.76) |
| **IMD quintile** |  |  |  |  |  |  |  |  |
| 1 (most deprived) | 1337 (18.9) | 6.50 (6.16 - 6.85) | 2352 (23.1) | 6.54 (6.27 - 6.80) | 1743 (20.6) | 4.01 (3.82 - 4.20) | 1967 (21.9) | 6.13 (5.86 - 6.40) |
| 2 | 1043 (20.7) | 6.69 (6.28 - 7.10) | 1645 (24.1) | 7.35 (7.00 - 7.70) | 1449 (19.3) | 4.18 (3.96 - 4.39) | 1673 (25.2) | 7.16 (6.82 - 7.50) |
| 3 | 896 (22.6) | 7.22 (6.75 - 7.69) | 1379 (25.3) | 7.74 (7.33 - 8.14) | 1330 (19.1) | 4.05 (3.83 - 4.26) | 1512 (26.2) | 7.51 (7.13 - 7.88) |
| 4 | 756 (23.1) | 7.29 (6.77 - 7.81) | 1057 (24.5) | 7.71 (7.24 - 8.17) | 1183 (19.9) | 4.24 (3.99 - 4.48) | 1288 (27.6) | 8.08 (7.64 - 8.52) |
| 5 (least deprived) | 574 (21.5) | 6.40 (5.88 - 6.93) | 798 (23.5) | 7.51 (6.99 - 8.03) | 952 (19.9) | 4.22 (3.95 - 4.49) | 1015 (28.0) | 8.25 (7.75 - 8.76) |

**Supplementary Table 4.** Crude and adjusted HRs for receipt of first fit note for comparisons of hospitalised cohorts, overall and by follow-up time post-diagnosis.

| Time period | Fit note rate per 100 person-months (95%CI) |  | Crude HR (95%CI) | Age-sex adjusted HR (95%CI) | Fully adjusted HR* (95%CI) |
| --- | --- | --- | --- | --- | --- |
|  | COVID-19 cohort | Comparator cohort |  |  |  |
| **2020 COVID-19 cohort vs 2019 pneumonia cohort** |  |  |  |  |  |
| 0-30 days | 18.84 (18.18 - 19.49) | 22.06 (21.46 - 22.66) | 0.85 (0.81 - 0.88) | 0.83 (0.78 - 0.88) | 0.72 (0.68 - 0.77) |
| 0-90 days | 11.44 (11.09 - 11.79) | 12.47 (12.17 - 12.76) | 0.86 (0.83 - 0.89) | 0.83 (0.78 - 0.88) | 0.74 (0.69 - 0.78) |
| 0-150 days | 8.66 (8.40 - 8.92) | 9.28 (9.06 - 9.49) | 0.88 (0.85 - 0.92) | 0.84 (0.79 - 0.88) | 0.75 (0.71 - 0.80) |
| Overall | 6.78 (6.59 - 6.98) | 7.17 (7.01 - 7.34) | 0.90 (0.86 - 0.93) | 0.85 (0.80 - 0.89) | 0.77 (0.73 - 0.81) |
| **2021 COVID-19 cohort vs 2019 pneumonia cohort** |  |  |  |  |  |
| 0-30 days | 21.39 (20.81 - 21.96) | 22.06 (21.46 - 22.66) | 0.97 (0.93 - 1.00) | 0.98 (0.92 - 1.03) | 0.83 (0.78 - 0.88) |
| 0-90 days | 11.65 (11.37 - 11.93) | 12.47 (12.17 - 12.76) | 0.93 (0.90 - 0.96) | 0.92 (0.88 - 0.97) | 0.81 (0.77 - 0.85) |
| 0-150 days | 9.03 (8.82 - 9.24) | 9.28 (9.06 - 9.49) | 0.93 (0.90 - 0.96) | 0.91 (0.87 - 0.96) | 0.80 (0.77 - 0.85) |
| Overall | 7.19 (7.03 - 7.36) | 7.17 (7.01 - 7.34) | 0.94 (0.91 - 0.97) | 0.92 (0.87 - 0.96) | 0.81 (0.77 - 0.86) |
| **2022 COVID-19 cohort vs 2019 pneumonia cohort** |  |  |  |  |  |
| 0-30 days | 12.92 (12.52 - 13.32) | 22.06 (21.46 - 22.66) | 0.59 (0.57 - 0.62) | 0.58 (0.54 - 0.61) | 0.56 (0.53 - 0.60) |
| 0-90 days | 7.05 (6.87 - 7.24) | 12.47 (12.17 - 12.75) | 0.61 (0.59 - 0.62) | 0.59 (0.56 - 0.62) | 0.58 (0.56 - 0.61) |
| 0-150 days | 5.22 (5.09 - 5.35) | 9.28 (9.06 - 9.49) | 0.63 (0.61 - 0.65) | 0.61 (0.58 - 0.64) | 0.60 (0.58 - 0.63) |
| Overall | 4.13 (4.03 - 4.22) | 7.17 (7.01 - 7.34) | 0.65 (0.63 - 0.67) | 0.63 (0.60 - 0.66) | 0.62 (0.59 - 0.65) |

\*Fully adjusted models include age, sex, IMD quintile, region, ethnicity, obesity, smoking status, hypertension, diabetes, chronic respiratory disease, asthma, chronic cardiac disease, lung cancer, haematological cancer, other cancer, chronic liver disease, other neurological disease, organ transplant, asplenia, HIV, permanent immunodeficiency, and rheumatoid arthritis/systemic lupus erythematosus/psoriasis.

**Supplementary Table 5.**
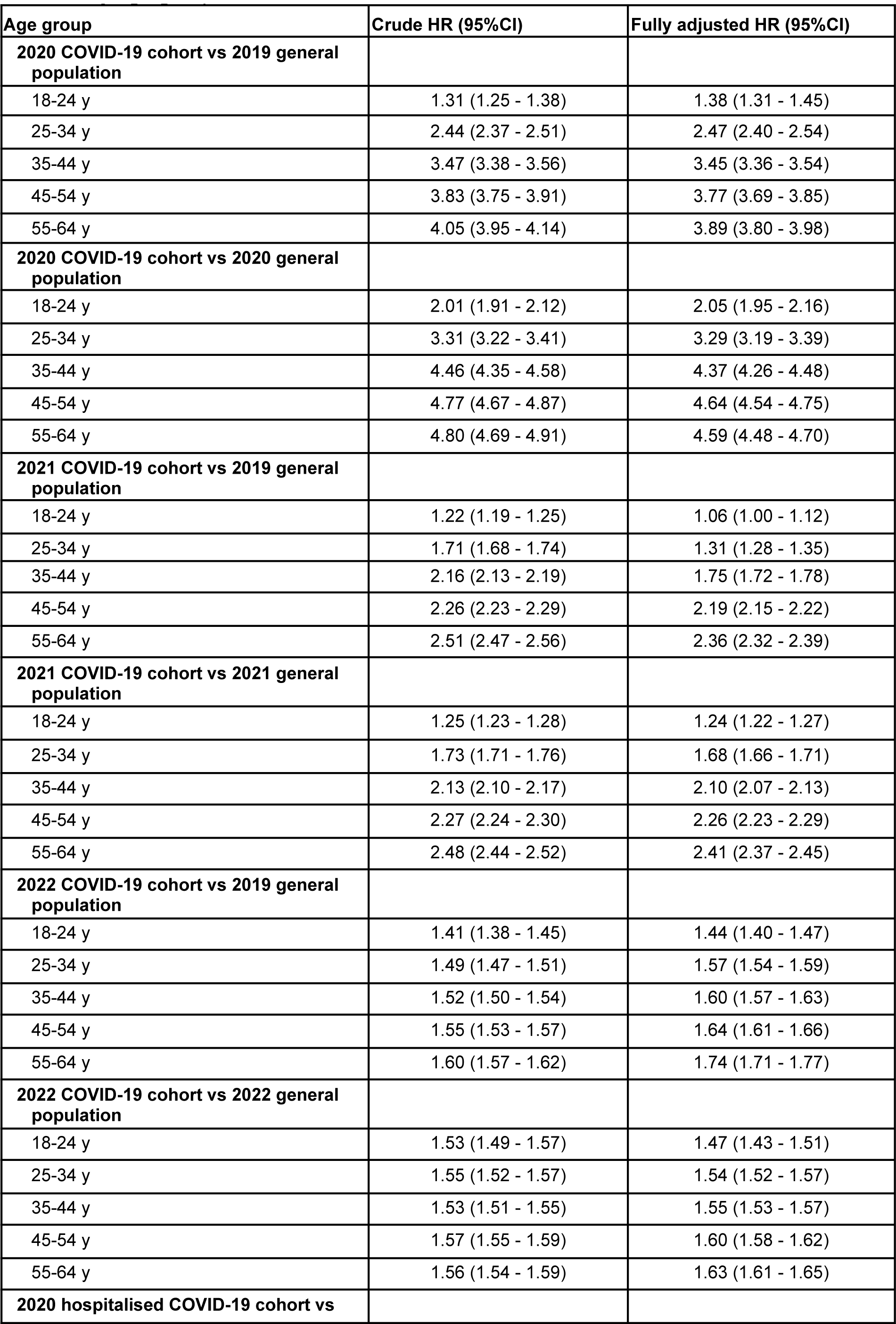
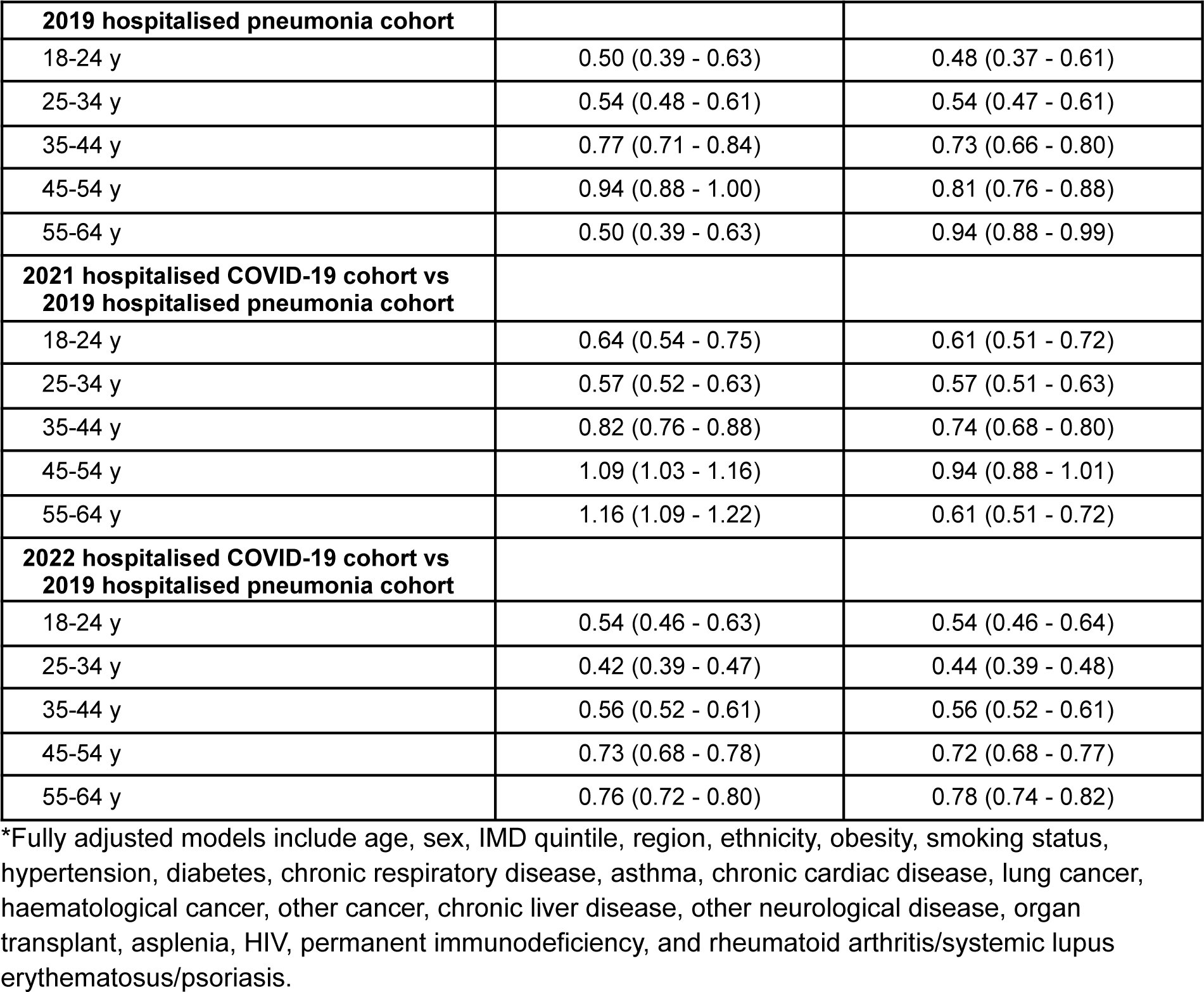
Crude and adjusted hazard ratio for first fit note for all comparisons, stratified by age group.

**Supplementary Table 6.** Crude and adjusted hazard ratio of first fit note for all comparisons, stratified by sex.

| Sex | Crude HR (95%CI) | Fully adjusted HR (95%CI) |
| --- | --- | --- |
| **2020 COVID-19 cohort vs 2022 general population** |  |  |
| Female | 3.39 (3.34 - 3.44) | 3.40 (3.35 - 3.45) |
| Male | 2.86 (2.80 - 2.92) | 2.84 (2.78 - 2.89) |
| **2020 COVID-19 cohort vs 2020 general population** |  |  |
| Female | 4.22 (4.16 - 4.29) | 4.22 (4.15 - 4.28) |
| Male | 3.91 (3.83 - 4.00) | 3.79 (3.71 - 3.87) |
| **2021 COVID-19 cohort vs 2019 general population** |  |  |
| Female | 2.08 (2.06 - 2.10) | 2.08 (2.06 - 2.10) |
| Male | 1.84 (1.82 - 1.86) | 1.93 (1.90 - 1.95) |
| **2021 COVID-19 cohort vs 2021 general population** |  |  |
| Female | 2.01 (1.99 - 2.03) | 2.01 (2.00 - 2.03) |
| Male | 1.93 (1.91 - 1.95) | 1.95 (1.93 - 1.98) |
| **2022 COVID-19 cohort vs 2019 general population** |  |  |
| Female | 1.54 (1.52 - 1.55) | 1.52 (1.51 - 1.54) |
| Male | 1.50 (1.48 - 1.52) | 1.61 (1.59 - 1.64) |
| **2022 COVID-19 cohort vs 2022 general population** |  |  |
| Female | 1.53 (1.51 - 1.54) | 1.53 (1.52 - 1.54) |
| Male | 1.60 (1.58 - 1.62) | 1.67 (1.65 - 1.69) |
| **2029 hospitalised COVID-19 cohort vs 2019 hospitalised pneumonia cohort** |  |  |
| Female | 0.95 (0.91 - 1.01) | 0.78 (0.72 - 0.85) |
| Male | 0.85 (0.81 - 0.89) | 0.75 (0.69 - 0.81) |
| **2021 hospitalised COVID-19 cohort vs 2019 hospitalised pneumonia cohort** |  |  |
| Female | 0.87 (0.83 - 0.92) | 0.76 (0.71 - 0.82) |
| Male | 1.02 (0.97 - 1.06) | 0.88 (0.82 - 0.94) |
| **2022 hospitalised COVID-19 cohort vs 2019 hospitalised pneumonia cohort** |  |  |
| Female | 0.61 (0.59 - 0.64) | 0.57 (0.53 - 0.61) |
| Male | 0.70 (0.67 - 0.74) | 0.70 (0.66 - 0.75) |

\*Fully adjusted models include age, sex, IMD quintile, region, ethnicity, obesity, smoking status, hypertension, diabetes, chronic respiratory disease, asthma, chronic cardiac disease, lung cancer, haematological cancer, other cancer, chronic liver disease, other neurological disease, organ transplant, asplenia, HIV, permanent immunodeficiency, and rheumatoid arthritis/systemic lupus erythematosus/psoriasis.

**Supplementary Table 7.** Crude and adjusted hazard ratio of first fit note for all comparisons, stratified by ethnicity.

| **Ethnicity** | **Crude HR (95%CI)** | **Fully adjusted HR (95%CI)** |
| --- | --- | --- |
| **2020 COVID-19 cohort vs 2019 general population** |  |  |
| White | 3.13 (3.08 - 3.18) | 3.13 (3.08 - 3.18) |
| Asian or Asian British | 3.71 (3.59 - 3.84) | 3.60 (3.48 - 3.73) |
| Black | 3.81 (3.56 - 4.08) | 3.76 (3.50 - 4.03) |
| Mixed | 3.04 (2.73 - 3.38) | 2.99 (2.68 - 3.33) |
| Other | 5.33 (4.83 - 5.89) | 4.54 (4.08 - 5.05) |
| Not stated | 3.08 (3.00 - 3.15) | 3.02 (2.95 - 3.10) |
| **2020 COVID-19 cohort vs 2020 general population** |  |  |
| White | 4.05 (3.99 - 4.11) | 4.01 (3.95 - 4.08) |
| Asian or Asian British | 4.44 (4.30 - 4.60) | 4.31 (4.16 - 4.47) |
| Black | 5.12 (4.78 - 5.49) | 4.95 (4.60 - 5.32) |
| Mixed | 4.00 (3.59 - 4.45) | 3.89 (3.48 - 4.34) |
| Other | 6.74 (6.09 - 7.46) | 5.89 (5.28 - 6.56) |
| Not stated | 4.02 (3.91 - 4.12) | 3.91 (3.81 - 4.01) |
| **2021 COVID-19 cohort vs 2019 general population** |  |  |
| White | 1.89 (1.87 - 1.91) | 1.94 (1.92 - 1.96) |
| Asian or Asian British | 2.84 (2.76 - 2.92) | 2.93 (2.84 - 3.02) |
| Black | 2.59 (2.47 - 2.72) | 2.88 (2.73 - 3.04) |
| Mixed | 2.01 (1.87 - 2.15) | 2.14 (1.98 - 2.30) |
| Other | 3.42 (3.18 - 3.67) | 3.22 (2.97 - 3.49) |
| Not stated | 1.82 (1.79 - 1.85) | 1.91 (1.88 - 1.94) |
| **2021 COVID-19 cohort vs 2021 general population** |  |  |
| White | 1.91 (1.90 - 1.93) | 1.94 (1.92 - 1.96) |
| Asian or Asian British | 2.69 (2.62 - 2.76) | 2.62 (2.55 - 2.70) |
| Black | 2.54 (2.42 - 2.66) | 2.62 (2.50 - 2.74) |
| Mixed | 2.09 (1.95 - 2.23) | 2.08 (1.95 - 2.23) |
| Other | 3.23 (3.03 - 3.45) | 2.90 (2.71 - 3.11) |
| Not stated | 1.84 (1.81 - 1.87) | 1.87 (1.84 - 1.90) |
| **2022 COVID-19 cohort vs 2019 general population** |  |  |
| White | 1.46 (1.45 - 1.47) | 1.48 (1.47 - 1.50) |
| Asian or Asian British | 2.21 (2.15 - 2.28) | 2.48 (2.40 - 2.58) |
| Black | 1.94 (1.85 - 2.04) | 1.88 (1.77 - 1.99) |
| Mixed | 1.57 (1.47 - 1.68) | 1.66 (1.54 - 1.79) |
| Other | 2.32 (2.16 - 2.48) | 2.23 (2.06 - 2.42) |
| Not stated | 1.62 (1.59 - 1.64) | 1.54 (1.51 - 1.57) |
| **2022 COVID-19 cohort vs 2022 general population** |  |  |
| White | 1.49 (1.48 - 1.50) | 2.10 (2.04 - 2.16) |
| Asian or Asian British | 1.97 (1.91 - 2.02) | 1.69 (1.61 - 1.77) |
| Black | 1.71 (1.64 - 1.80) | 1.60 (1.51 - 1.71) |
| Mixed | 1.54 (1.45 - 1.64) | 1.98 (1.86 - 2.10) |
| Other | 1.93 (1.82 - 2.04) | 1.61 (1.59 - 1.63) |
| Not stated | 1.63 (1.61 - 1.66) | 2.10 (2.04 - 2.16) |
| **2020 hospitalised COVID-19 cohort vs 2019 hospitalised pneumonia cohort** |  |  |
| White | 0.93 (0.89 - 0.97) | 0.78 (0.72 - 0.83) |
| Asian or Asian British | 0.98 (0.86 - 1.12) | 0.90 (0.76 - 1.08) |
| Black | 0.97 (0.79 - 1.19) | 0.80 (0.60 - 1.07) |
| Mixed | 0.96 (0.68 - 1.35) | 0.58 (0.35 - 0.94) |
| Other | 0.83 (0.61 - 1.14) | 0.77 (0.51 - 1.18) |
| Not stated | 0.83 (0.77 - 0.90) | 0.74 (0.65 - 0.83) |
| **2021 hospitalised COVID-19 cohort vs 2019 hospitalised pneumonia cohort** |  |  |
| White | 0.99 (0.95 - 1.03) | 0.81 (0.76 - 0.87) |
| Asian or Asian British | 0.94 (0.83 - 1.07) | 1.02 (0.86 - 1.22) |
| Black | 1.03 (0.85 - 1.25) | 0.90 (0.69 - 1.17) |
| Mixed | 0.83 (0.60 - 1.14) | 0.55 (0.35 - 0.84) |
| Other | 0.70 (0.52 - 0.96) | 0.73 (0.47 - 1.12) |
| Not stated | 0.88 (0.82 - 0.94) | 0.76 (0.69 - 0.85) |
| **2022 hospitalised COVID-19 cohort vs 2019 hospitalised pneumonia cohort** |  |  |
| White | 0.65 (0.63 - 0.68) | 0.62 (0.58 - 0.65) |
| Asian or Asian British | 0.75 (0.65 - 0.86) | 0.75 (0.63 - 0.90) |
| Black | 0.72 (0.58 - 0.90) | 0.68 (0.52 - 0.90) |
| Mixed | 0.62 (0.44 - 0.88) | 0.51 (0.32 - 0.80) |
| Other | 0.58 (0.42 - 0.80) | 0.61 (0.41 - 0.91) |
| Not stated | 0.61 (0.57 - 0.66) | 0.60 (0.54 - 0.66) |

\*Fully adjusted models include age, sex, IMD quintile, region, ethnicity, obesity, smoking status, hypertension, diabetes, chronic respiratory disease, asthma, chronic cardiac disease, lung cancer, haematological cancer, other cancer, chronic liver disease, other neurological disease, organ transplant, asplenia, HIV, permanent immunodeficiency, and rheumatoid arthritis/systemic lupus erythematosus/psoriasis.

**Supplementary Table 8.** Crude and adjusted hazard ratio of first fit note for all comparisons, stratified by IMD quintile.

| IMD quintile | Crude HR (95%CI) | Fully adjusted HR (95%CI) |
| --- | --- | --- |
| **2020 COVID-19 cohort vs 2019 general population** |  |  |
| 1 (most deprived) | 2.64 (2.59 - 2.70) | 2.70 (2.64 - 2.76) |
| 2 | 3.05 (2.98 - 3.13) | 3.06 (2.98 - 3.14) |
| 3 | 3.45 (3.36 - 3.54) | 3.43 (3.33 - 3.52) |
| 4 | 3.71 (3.61 - 3.82) | 3.71 (3.61 - 3.82) |
| 5 (least deprived) | 3.86 (3.74 - 3.99) | 3.93 (3.80 - 4.06) |
| **2020 COVID-19 cohort vs 2020 general population** |  |  |
| 1 (most deprived) | 3.44 (3.36 - 3.51) | 3.41 (3.33 - 3.48) |
| 2 | 3.96 (3.86 - 4.06) | 3.91 (3.81 - 4.02) |
| 3 | 4.41 (4.29 - 4.53) | 4.36 (4.24 - 4.49) |
| 4 | 4.72 (4.59 - 4.86) | 4.71 (4.57 - 4.86) |
| 5 (least deprived) | 5.02 (4.86 - 5.18) | 5.15 (4.98 - 5.32) |
| **2021 COVID-19 cohort vs 2019 general population** |  |  |
| 1 (most deprived) | 1.84 (1.81 - 1.86) | 1.86 (1.84 - 1.89) |
| 2 | 1.93 (1.90 - 1.96) | 1.98 (1.94 - 2.01) |
| 3 | 2.04 (2.00 - 2.07) | 2.10 (2.06 - 2.14) |
| 4 | 2.09 (2.05 - 2.13) | 2.15 (2.10 - 2.19) |
| 5 (least deprived) | 2.18 (2.14 - 2.23) | 2.25 (2.20 - 2.30) |
| **2021 COVID-19 cohort vs 2021 general population** |  |  |
| 1 (most deprived) | 1.77 (1.75 - 1.79) | 1.77 (1.74 - 1.79) |
| 2 | 1.93 (1.90 - 1.96) | 1.95 (1.92 - 1.98) |
| 3 | 2.06 (2.02 - 2.09) | 2.09 (2.06 - 2.13) |
| 4 | 2.09 (2.05 - 2.12) | 2.15 (2.11 - 2.19) |
| 5 (least deprived) | 2.21 (2.17 - 2.26) | 2.30 (2.26 - 2.35) |
| **2022 COVID-19 cohort vs 2019 general population** |  |  |
| 1 (most deprived) | 1.68 (1.66 - 1.70) | 1.59 (1.57 - 1.62) |
| 2 | 1.64 (1.62 - 1.67) | 1.57 (1.54 - 1.59) |
| 3 | 1.62 (1.60 - 1.65) | 1.55 (1.52 - 1.58) |
| 4 | 1.64 (1.62 - 1.67) | 1.53 (1.49 - 1.56) |
| 5 (least deprived) | 1.62 (1.59 - 1.65) | 1.50 (1.47 - 1.54) |
| **2022 COVID-19 cohort vs 2022 general population** |  |  |
| 1 (most deprived) | 1.58 (1.55 - 1.60) | 1.55 (1.53 - 1.57) |
| 2 | 1.60 (1.58 - 1.62) | 1.59 (1.56 - 1.61) |
| 3 | 1.58 (1.56 - 1.61) | 1.57 (1.55 - 1.59) |
| 4 | 1.61 (1.59 - 1.64) | 1.60 (1.58 - 1.63) |
| 5 (least deprived) | 1.56 (1.54 - 1.59) | 1.56 (1.53 - 1.58) |
| **2020 hospitalised COVID-19 cohort vs 2019 hospitalised pneumonia cohort** |  |  |
| 1 (most deprived) | 0.96 (0.89 - 1.03) | 0.76 (0.68 - 0.84) |
| 2 | 0.9 (0.83 - 0.97) | 0.73 (0.65 - 0.82) |
| 3 | 0.93 (0.85 - 1.01) | 0.83 (0.73 - 0.94) |
| 4 | 0.88 (0.81 - 0.96) | 0.84 (0.73 - 0.96) |
| 5 (least deprived) | 0.78 (0.70 - 0.86) | 0.71 (0.60 - 0.83) |
| **2021 hospitalised COVID-19 cohort vs 2019 hospitalised pneumonia cohort** |  |  |
| 1 (most deprived) | 1.02 (0.97 - 1.09) | 0.85 (0.78 - 0.93) |
| 2 | 0.96 (0.90 - 1.03) | 0.80 (0.72 - 0.89) |
| 3 | 0.96 (0.90 - 1.04) | 0.89 (0.79 - 0.99) |
| 4 | 0.88 (0.81 - 0.96) | 0.79 (0.70 - 0.90) |
| 5 (least deprived) | 0.82 (0.75 - 0.90) | 0.69 (0.60 - 0.79) |
| **2022 hospitalised COVID-19 cohort vs 2019 hospitalised pneumonia cohort** |  |  |
| 1 (most deprived) | 0.71 (0.67 - 0.76) | 0.66 (0.60 - 0.72) |
| 2 | 0.65 (0.61 - 0.70) | 0.63 (0.57 - 0.70) |
| 3 | 0.62 (0.57 - 0.66) | 0.63 (0.57 - 0.70) |
| 4 | 0.60 (0.56 - 0.65) | 0.57 (0.51 - 0.64) |
| 5 (least deprived) | 0.60 (0.55 - 0.65) | 0.59 (0.52 - 0.67) |

\*Fully adjusted models include age, sex, IMD quintile, region, ethnicity, obesity, smoking status, hypertension, diabetes, chronic respiratory disease, asthma, chronic cardiac disease, lung cancer, haematological cancer, other cancer, chronic liver disease, other neurological disease, organ transplant, asplenia, HIV, permanent immunodeficiency, and rheumatoid arthritis/systemic lupus erythematosus/psoriasis.

**Supplementary Table 9.** Crude and adjusted hazard ratio of first fit note for all comparisons, stratified by region.

| Region | Crude HR (95%CI) | Fully adjusted HR (95%CI) |
| --- | --- | --- |
| **2020 COVID-19 cohort vs 2019 general population** |  |  |
| East Midlands | 3.13 (3.05 - 3.22) | 3.12 (3.04 - 3.21) |
| East | 3.34 (3.24 - 3.45) | 3.17 (3.07 - 3.27) |
| London | 3.68 (3.45 - 3.92) | 3.24 (3.03 - 3.47) |
| North East | 3.12 (3.00 - 3.24) | 3.15 (3.03 - 3.28) |
| North West | 3.20 (3.11 - 3.30) | 3.24 (3.14 - 3.33) |
| South East | 3.28 (3.06 - 3.52) | 3.14 (2.92 - 3.37) |
| South West | 3.44 (3.29 - 3.61) | 3.36 (3.21 - 3.52) |
| West Midlands | 3.29 (3.14 - 3.46) | 3.32 (3.16 - 3.49) |
| Yorkshire & The Humber | 3.15 (3.07 - 3.22) | 3.16 (3.08 - 3.24) |
| **2020 COVID-19 cohort vs 2029 general population** |  |  |
| East Midlands | 4.05 (3.94 - 4.16) | 3.98 (3.87 - 4.09) |
| East | 4.56 (4.42 - 4.71) | 4.28 (4.14 - 4.43) |
| London | 5.22 (4.89 - 5.58) | 4.57 (4.26 - 4.89) |
| North East | 3.86 (3.71 - 4.02) | 3.86 (3.71 - 4.02) |
| North West | 3.94 (3.82 - 4.06) | 3.92 (3.80 - 4.04) |
| South East | 4.34 (4.04 - 4.65) | 4.15 (3.86 - 4.46) |
| South West | 4.55 (4.35 - 4.77) | 4.40 (4.19 - 4.61) |
| West Midlands | 4.37 (4.16 - 4.59) | 4.34 (4.13 - 4.57) |
| Yorkshire & The Humber | 4.02 (3.92 - 4.12) | 3.97 (3.88 - 4.07) |
| **2021 COVID-19 cohort vs 2019 general population** |  |  |
| East Midlands | 2.05 (2.02 - 2.09) | 2.06 (2.02 - 2.10) |
| East | 2.01 (1.97 - 2.05) | 1.97 (1.93 - 2.01) |
| London | 2.63 (2.53 - 2.74) | 2.62 (2.50 - 2.74) |
| North East | 1.96 (1.91 - 2.01) | 1.97 (1.91 - 2.03) |
| North West | 1.85 (1.81 - 1.88) | 1.87 (1.83 - 1.91) |
| South East | 2.02 (1.94 - 2.10) | 1.98 (1.90 - 2.07) |
| South West | 2.03 (1.98 - 2.09) | 1.99 (1.93 - 2.04) |
| West Midlands | 2.10 (2.03 - 2.17) | 2.18 (2.1 - 2.26) |
| Yorkshire & The Humber | 2.03 (1.99 - 2.06) | 2.04 (2.00 - 2.07) |
| **2021 COVID-19 cohort vs 2021 general population** |  |  |
| East Midlands | 1.95 (1.92 - 1.98) | 1.99 (1.96 - 2.02) |
| East | 2.07 (2.03 - 2.11) | 2.07 (2.04 - 2.11) |
| London | 2.69 (2.59 - 2.79) | 2.60 (2.50 - 2.70) |
| North East | 1.85 (1.80 - 1.90) | 1.89 (1.84 - 1.94) |
| North West | 1.73 (1.69 - 1.76) | 1.76 (1.72 - 1.79) |
| South East | 2.02 (1.95 - 2.09) | 2.05 (1.98 - 2.12) |
| South West | 2.02 (1.97 - 2.06) | 2.06 (2.02 - 2.11) |
| West Midlands | 2.04 (1.98 - 2.11) | 2.08 (2.01 - 2.14) |
| Yorkshire & The Humber | 1.92 (1.89 - 1.95) | 1.95 (1.92 - 1.99) |
| **2022 COVID-19 cohort vs 2019 general population** |  |  |
| East Midlands | 1.71 (1.68 - 1.73) | 1.58 (1.55 - 1.61) |
| East | 1.61 (1.58 - 1.64) | 1.50 (1.47 - 1.53) |
| London | 1.91 (1.84 - 1.98) | 1.96 (1.88 - 2.05) |
| North East | 1.68 (1.63 - 1.73) | 1.56 (1.51 - 1.61) |
| North West | 1.63 (1.60 - 1.66) | 1.49 (1.46 - 1.53) |
| South East | 1.67 (1.61 - 1.73) | 1.53 (1.47 - 1.59) |
| South West | 1.66 (1.62 - 1.69) | 1.47 (1.44 - 1.51) |
| West Midlands | 1.80 (1.75 - 1.86) | 1.74 (1.68 - 1.81) |
| Yorkshire & The Humber | 1.65 (1.63 - 1.68) | 1.56 (1.53 - 1.59) |
| **2022 COVID-19 cohort vs 2022 general population** |  |  |
| East Midlands | 1.55 (1.53 - 1.58) | 1.57 (1.55 - 1.60) |
| East | 1.55 (1.53 - 1.58) | 1.57 (1.55 - 1.59) |
| London | 1.79 (1.74 - 1.86) | 1.90 (1.83 - 1.96) |
| North East | 1.50 (1.46 - 1.54) | 1.53 (1.49 - 1.57) |
| North West | 1.51 (1.48 - 1.54) | 1.51 (1.48 - 1.54) |
| South East | 1.54 (1.50 - 1.58) | 1.54 (1.50 - 1.58) |
| South West | 1.54 (1.52 - 1.57) | 1.54 (1.52 - 1.57) |
| West Midlands | 1.62 (1.57 - 1.67) | 1.65 (1.60 - 1.71) |
| Yorkshire & The Humber | 1.53 (1.51 - 1.56) | 1.58 (1.55 - 1.61) |
| **2020 hospitalised COVID-19 cohort vs 2019 hospitalised pneumonia cohort** |  |  |
| East Midlands | 0.83 (0.77 - 0.91) | 0.69 (0.61 - 0.78) |
| East | 0.78 (0.67 - 0.91) | 0.74 (0.65 - 0.84) |
| London | 0.94 (0.82 - 1.09) | 0.65 (0.51 - 0.83) |
| North East | 1.06 (0.95 - 1.18) | 0.81 (0.65 - 1.02) |
| North West | 0.75 (0.63 - 0.90) | 0.89 (0.76 - 1.04) |
| South East | 1.10 (0.98 - 1.23) | 0.58 (0.45 - 0.76) |
| South West | 0.88 (0.75 - 1.02) | 1.08 (0.91 - 1.28) |
| West Midlands | 0.96 (0.88 - 1.05) | 0.74 (0.59 - 0.93) |
| Yorkshire & The Humber | 0.83 (0.77 - 0.91) | 0.78 (0.69 - 0.90) |
| **2021 hospitalised COVID-19 cohort vs 2019 hospitalised pneumonia cohort** |  |  |
| East Midlands | 0.91 (0.85 - 0.98) | 0.78 (0.69 - 0.87) |
| East | 0.92 (0.85 - 0.99) | 0.79 (0.70 - 0.88) |
| London | 0.86 (0.74 - 1.00) | 0.80 (0.65 - 1.00) |
| North East | 0.94 (0.83 - 1.08) | 0.83 (0.68 - 1.03) |
| North West | 0.98 (0.89 - 1.09) | 0.81 (0.70 - 0.95) |
| South East | 0.90 (0.78 - 1.04) | 0.79 (0.63 - 0.99) |
| South West | 0.99 (0.90 - 1.09) | 0.82 (0.71 - 0.95) |
| West Midlands | 0.96 (0.84 - 1.11) | 0.92 (0.74 - 1.15) |
| Yorkshire & The Humber | 0.98 (0.91 - 1.06) | 0.86 (0.77 - 0.97) |
| **2022 hospitalised COVID-19 cohort vs 2019 hospitalised pneumonia cohort** |  |  |
| East Midlands | 0.68 (0.63 - 0.73) | 0.68 (0.61 - 0.75) |
| East | 0.60 (0.56 - 0.65) | 0.60 (0.54 - 0.66) |
| London | 0.61 (0.52 - 0.71) | 0.72 (0.58 - 0.90) |
| North East | 0.59 (0.52 - 0.68) | 0.58 (0.48 - 0.70) |
| North West | 0.69 (0.63 - 0.77) | 0.61 (0.53 - 0.71) |
| South East | 0.58 (0.51 - 0.66) | 0.56 (0.46 - 0.67) |
| South West | 0.63 (0.58 - 0.69) | 0.58 (0.51 - 0.67) |
| West Midlands | 0.74 (0.64 - 0.86) | 0.72 (0.58 - 0.88) |
| Yorkshire & The Humber | 0.69 (0.64 - 0.75) | 0.61 (0.54 - 0.68) |

\*Fully adjusted models include age, sex, IMD quintile, region, ethnicity, obesity, smoking status, hypertension, diabetes, chronic respiratory disease, asthma, chronic cardiac disease, lung cancer, haematological cancer, other cancer, chronic liver disease, other neurological disease, organ transplant, asplenia, HIV, permanent immunodeficiency, and rheumatoid arthritis/systemic lupus erythematosus/psoriasis.

**Supplementary Figure 3.**
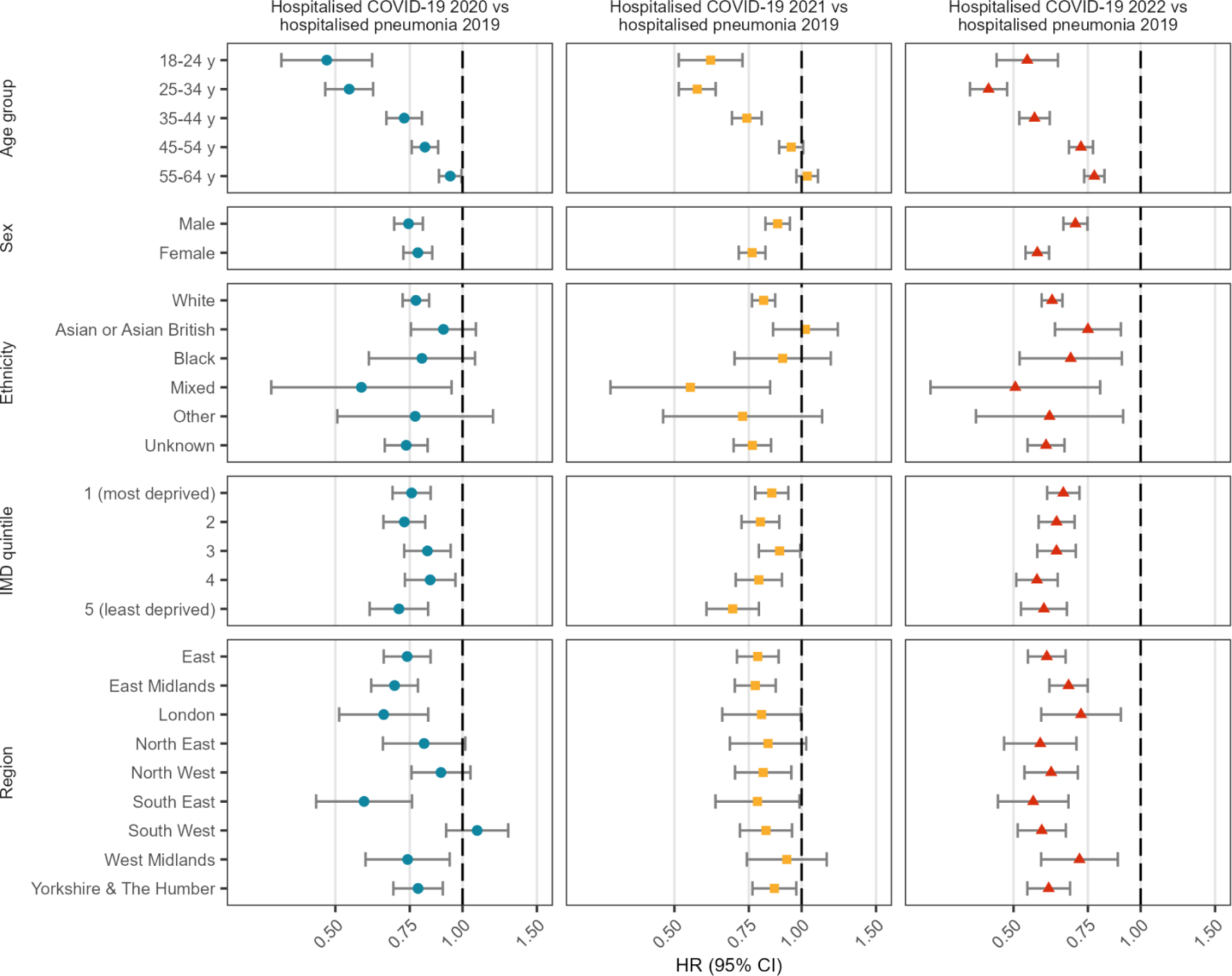
Adjusted hazard ratio of first fit note comparing hospitalised COVID-19 cohorts to people hospitalised with pneumonia in 2019, stratified by demographic categories and year *All models are adjusted for age, sex, ethnicity, IMD quintile and region, excluding the stratification variable. Models are additionally adjusted for obesity, smoking status, hypertension, diabetes, chronic respiratory disease, asthma, chronic cardiac disease, lung cancer, haematological cancer, other cancer, chronic liver disease, other neurological disease, organ transplant, asplenia, HIV, permanent immunodeficiency, and rheumatoid arthritis/systemic lupus erythematosus/psoriasis

## References

1. NHS Digital. Fit Notes Issued by GP Practices, England, September 2022 [Internet]. [cited 2023 Mar 6]. Available from: https://digital.nhs.uk/data-and-information/publications/statistical/fit-notes-issued-by-gp-practices/september-2022

2. Dorrington S, Roberts E, Mykletun A, Hatch S, Madan I, Hotopf M. Systematic review of fit note use for workers in the UK. Occup Environ Med. 2018 Jul 1;75(7):530–9.

3. Henderson M, Clark C, Stansfeld S, Hotopf M. A lifecourse approach to long-term sickness absence--a cohort study. PloS One. 2012;7(5):e36645.

4. UK Department for Work and Pensions. An evaluation of the Statement of Fitness for Work (fit note): Survey of employees (RR 840) [Internet]. [cited 2023 Mar 6]. Report No.: 840. Available from: https://www.gov.uk/government/publications/an-evaluation-of-the-statement-of-fitness-for-work-fit-note-survey-of-employees-rr-840

5. Office for National Statistics. Coronavirus (COVID-19) Infection Survey technical article: Cumulative incidence of the percentage of people who have been infected with COVID-19 by variant and age, England [Internet]. 2023 [cited 2023 Jul 6]. Available from: https://www.ons.gov.uk/peoplepopulationandcommunity/healthandsocialcare/conditionsanddiseases/articles/coronaviruscovid19infectionsurveytechnicalarticlecumulativeincidenceofthenumberofpeoplewhohavebeeninfectedwithcovid19byvariantandageengland/9february2023

6. Office for National Statistics. Cumulative COVID-19 infections time series, England [Internet]. 2023 [cited 2023 Jul 6]. Available from: https://www.ons.gov.uk/peoplepopulationandcommunity/healthandsocialcare/conditionsanddiseases/datasets/cumulativecovid19infectionstimeseriesengland

7. Office for National Statistics. Prevalence of ongoing symptoms following coronavirus (COVID-19) infection in the UK [Internet]. 2023 [cited 2023 Mar 31]. Available from: https://www.ons.gov.uk/peoplepopulationandcommunity/healthandsocialcare/conditionsanddiseases/bulletins/prevalenceofongoingsymptomsfollowingcoronaviruscovid19infectionintheuk/latest

8. Dorrington S, Carr E, Polling C, Stevelink S, Ashworth M, Roberts E, et al. Health condition at first fit note and number of fit notes: a longitudinal study of primary care records in south London. BMJ Open. 2021 Mar 1;11(3):e043889.

9. Michelen M, Manoharan L, Elkheir N, Cheng V, Dagens A, Hastie C, et al. Characterising long COVID: a living systematic review. BMJ Glob Health. 2021 Sep 1;6(9):e005427.

10. Davis HE, McCorkell L, Vogel JM, Topol EJ. Long COVID: major findings, mechanisms and recommendations. Nat Rev Microbiol. 2023 Mar;21(3):133–46.

11. Walker AJ, MacKenna B, Inglesby P, Tomlinson L, Rentsch CT, Curtis HJ, et al. Clinical coding of long COVID in English primary care: a federated analysis of 58 million patient records in situ using OpenSAFELY. Br J Gen Pract. 2021 Nov 1;71(712):e806–14.

12. Hernán MA. The Hazards of Hazard Ratios. Epidemiol Camb Mass. 2010 Jan;21(1):13–5.

13. Andrews C, Schultze A, Curtis H, Hulme W, Tazare J, Evans S, et al. OpenSAFELY: Representativeness of electronic health record platform OpenSAFELY-TPP data compared to the population of England. Wellcome Open Res. 2022;7:191.

14. Griffith GJ, Morris TT, Tudball MJ, Herbert A, Mancano G, Pike L, et al. Collider bias undermines our understanding of COVID-19 disease risk and severity. Nat Commun. 2020 Nov 12;11(1):5749.

15. Office for National Statistics. A05 SA: Employment, unemployment and economic inactivity by age group (seasonally adjusted) [Internet]. [cited 2023 May 17]. Available from: https://www.ons.gov.uk/employmentandlabourmarket/peopleinwork/employmentandemployeetypes/datasets/employmentunemploymentandeconomicinactivitybyagegroupseasonallyadjusteda05sa

16. Nab L, Parker EPK, Andrews CD, Hulme WJ, Fisher L, Morley J, et al. Changes in COVID-19-related mortality across key demographic and clinical subgroups in England from 2020 to 2022: a retrospective cohort study using the OpenSAFELY platform. Lancet Public Health. 2023 May 1;8(5):e364–77.

17. UK Health Security Agency. England Summary | Coronavirus (COVID-19) in the UK [Internet]. 2023 [cited 2023 Mar 6]. Available from: https://coronavirus.data.gov.uk

18. Office for National Statistics. Coronavirus (COVID-19) latest insights [Internet]. 2023 [cited 2023 Mar 31]. Available from: https://www.ons.gov.uk/peoplepopulationandcommunity/healthandsocialcare/conditionsanddiseases/articles/coronaviruscovid19latestinsights/infections

19. Edge R, van der Plaat DA, Parsons V, Coggon D, van Tongeren M, Muiry R, et al. Changing patterns of sickness absence among healthcare workers in England during the COVID-19 pandemic. J Public Health Oxf Engl. 2022 Mar 7;44(1):e42–50.

20. Kadambari S, Goldacre R, Morris E, Goldacre MJ, Pollard AJ. Indirect effects of the covid-19 pandemic on childhood infection in England: population based observational study. BMJ. 2022 Jan 12;376:e067519.

21. Coronavirus (COVID-19) Infection Survey, characteristics of people testing positive for COVID-19, UK - Office for National Statistics [Internet]. [cited 2023 May 17]. Available from: https://www.ons.gov.uk/peoplepopulationandcommunity/healthandsocialcare/conditionsanddiseases/bulletins/coronaviruscovid19infectionsurveycharacteristicsofpeopletestingpositiveforcovid19uk/21september2022

22. Abu-Raddad LJ, Chemaitelly H, Bertollini R, National Study Group for COVID-19 Epidemiology. Severity of SARS-CoV-2 Reinfections as Compared with Primary Infections. N Engl J Med. 2021 Dec 23;385(26):2487–9.

23. Mensah AA, Lacy J, Stowe J, Seghezzo G, Sachdeva R, Simmons R, et al. Disease severity during SARS-COV-2 reinfection: a nationwide study. J Infect. 2022 Apr;84(4):542–50.

24. Willyard C. Are repeat COVID infections dangerous? What the science says. Nature. 2023 Apr 26;616(7958):650–2.

25. Bowe B, Xie Y, Al-Aly Z. Acute and postacute sequelae associated with SARS-CoV-2 reinfection. Nat Med. 2022 Nov;28(11):2398–405.

26. Sudre CH, Murray B, Varsavsky T, Graham MS, Penfold RS, Bowyer RC, et al. Attributes and predictors of long COVID. Nat Med. 2021 Apr;27(4):626–31.

27. Whitaker M, Elliott J, Chadeau-Hyam M, Riley S, Darzi A, Cooke G, et al. Persistent COVID-19 symptoms in a community study of 606,434 people in England. Nat Commun. 2022 Apr 12;13(1):1957.

28. Thompson EJ, Williams DM, Walker AJ, Mitchell RE, Niedzwiedz CL, Yang TC, et al. Long COVID burden and risk factors in 10 UK longitudinal studies and electronic health records. Nat Commun. 2022 Jun 28;13(1):3528.

29. Palmer W, Rolewicz L. Nuffield Trust. 2023 [cited 2023 Jun 30]. All is not well: Sickness absence in the NHS in England. Available from: https://www.nuffieldtrust.org.uk/resource/all-is-not-well-sickness-absence-in-the-nhs-in-england

